# “I like being autistic”: Assessing the benefit of autistic-led psychoeducation for autistic children

**DOI:** 10.1101/2024.07.12.24310317

**Authors:** Sinéad L. Mullally, Cherice C. Edwards, Alice E. Wood, Sophie E. Connolly, Hannah Constable, Stuart Watson, Jacqui Rodgers

## Abstract

**Background:** Despite receiving autism diagnoses in early life, autistic children are not routinely supported to understand these diagnoses post-diagnostically (1). Consequently, they typically grow-up lacking an accurate understanding of what it means to be autistic on both a collective and individual level (2). Without this foundational knowledge, children’s understanding of autism is garnered from how others perceive their autism, resulting in an understanding of autism, and of themselves, that is inherently negative (3). This lack of appreciation of their own individual needs, also denies them the important self-understanding afforded by the diagnosis in the first instance, alongside the opportunity to effectively self-advocate for themselves when these needs go unmet.

**Aims:** Here we sought to directly assess the benefit of a pre-recorded, online autistic-led psychoeducation course about autism and the lived experience of being autistic (i.e., ‘NeuroBears’ https://www.pandasonline.org), for children’s understanding of autism and their autistic experiences, their feeling about being autistic, their communication with others about their autistic experiences, and their confidence to self-advocate for their needs.

**Methods:** Using a concurrent embedded mixed-methods, repeated-measures design, autistic children (aged 8-14 years), completed a bespoke questionnaire exploring the above topics, both before and after completing NeuroBears at home with a nominated safe adult. A total of 63 children (mean age=10.57 years) completed sufficient content to be included in the analysis.

**Results:** Significant benefit was observed across a range of areas, including a significant improvement in the children’s knowledge and understanding of being autistic and of their unique strengths and challenges, a significant rebalancing of how the children viewed being autistic, evidence of emerging positive autistic identities and a growing sense of belongingness, a significant change in the children’s abilities to communicate about being autistic, and evidence of strengthening self-advocacy skills.

**Conclusion:** Learning about autism in a neutral and non-stigmatizing manner, and presented through the lens of autistic lived experience, conferred numerous benefits on autistic children’s self-understanding, emergent autistic identity, sense of belonging, and on their communication/self-advocacy skills. Future work is needed to establish the downstream benefits on wellbeing and quality of life.

## 1. Introduction

Approximately 1% of children are diagnosed autistic globally (4). Autism, under the biomedical model, is viewed through a lens of pathology, characterised as a specific constellation of deficits, symptoms, and disturbances (5). Autism, according to this perspective, is inherently negative - a deviation of normal development grounded in brain disease (6) that represents an intrinsic barrier to a good life (7). Emanating from this vantage point is an academic literature littered with dehumanising and harmful rhetoric (8, 9). This rhetoric is now so normalised on a societal level that it is commonplace for autistic people to be viewed as existing “on the fringe of human normality” (10) (pp. 53) and where it is seemingly acceptable to explicitly question the value of autistic people to humanity (11).

In addition to propagating stigma, viewing autism through a lens of pathology motivates the development of interventions to cure the person who is suffering (12). Hence, a constellation of autism interventions has been developed to do just this (13). Despite enormous sums of public and private money spent on such interventions, they typically have little or no scientific evidence base (14) and are increasingly associated with enduring trauma (15). Given the potential risk to the individual of exposure to potentially harmful interventions, in addition to the harm likely caused to the individuals’ self-concept and self-esteem via the biomedical model’s deficit focused conceptualisation of autism, one could question the benefit of receiving an autism diagnosis in the first instance.

Seemingly paradoxical therefore is the observation that there continues to be a significant increase in the number of individuals diagnosed autistic. For instance, a 2021 UK population-based cohort study reported a 787% exponential increase in autism diagnoses between the years 1998 and 2018 (16). Moreover, receiving an autism diagnosis is often viewed as a positive step forward, as it permits for a reconfiguration of self and an appreciation of ones’ own individual needs (17). However, these benefits appear to be most readily actualised in the presence of greater satisfaction with one’s autistic identity (18, 19), greater autism pride (19), and a greater sense of autistic social identity (18, 20).

Positive personal and social autistic identities typically emerge when autism is viewed as a sociocultural identity alongside a clinical diagnosis (21), and when autism is reliably understood and viewed as a neutral difference rather than as a pathology (7). From this perspective, the many positive aspects of being autistic are recognised, alongside the real-world challenges (22). Equally important is the intersection between the individual strengths and challenges and the facilitating and hindering factors in any specific context (23). This represents a cultural shift away from the biomedical’s singularly deficit-focused conceptualisation; a shift that has emerged alongside increasing autistic self-advocacy and within an increasingly unified autistic community (12).

Hence, it appears crucial that after receiving an autism diagnosis, autistic individuals are supported to reliably understand autism in a neutral and non-stigmatizing manner. Currently autistic people are not routinely provided with any form of direct post-diagnostic support in the UK and frustration with this status quo is widely documented in autistic individuals, parents of autistic children, and the professionals providing these diagnoses (24, 25). Whilst some autistic adults find this form of post-diagnostic support independently, e.g., via social media autistic community groups (26), autistic children are much less likely to organically encounter such support. It is within this void that children’s autistic identities develop, often by assimilating how others perceive their autism into their emergent identities, with the perceptions of teachers and school peers being of particular importance (27). Concerningly, this appears to produce personal autistic identities that are inherently negative and (in the words of autistic school children) results in them perceiving themselves to be “retarded”, “a freak”, or as having a “bad brain” (27). Similarly, a metasynthesis of 17 qualitative studies (which included participants from age 5-21 years of age) found that experiences of autistic young people at school informed their sense-making processes about themselves, and that self-peer interactions at school typically resulted in them coming to understand their sense of self as being ‘different’ in a negative way; a sense that was then either accentuated or minimised by their treatment by teaching staff (3).

Many within the autistic community work tirelessly to counter such narratives and to ultimately facilitate the emergence of more positive autistic identities in new generations of autistic individuals. Recognising the benefit that access to such information could afford autistic children post-diagnosis, the psychoeducation course, NeuroBears, was developed. NeuroBears was created by autistic individuals, for, and in collaboration with, autistic children. It is primarily aimed at autistic children aged 8-14 years-old who are new to understanding their autism. It aims to educate and increase children’s understanding of their autistic experiences from a neutral viewpoint and to provide them with the context and language to describe and discuss these experiences with others. Ultimately, NeuroBears seeks to empower autistic children to build the confidence to self-advocate for their need and the knowledge to seek out safe spaces to be, whilst simultaneously promoting the development of a positive autistic identity and awareness of the wider autistic community to which they belong (see Figure 1).

**Figure 1.**
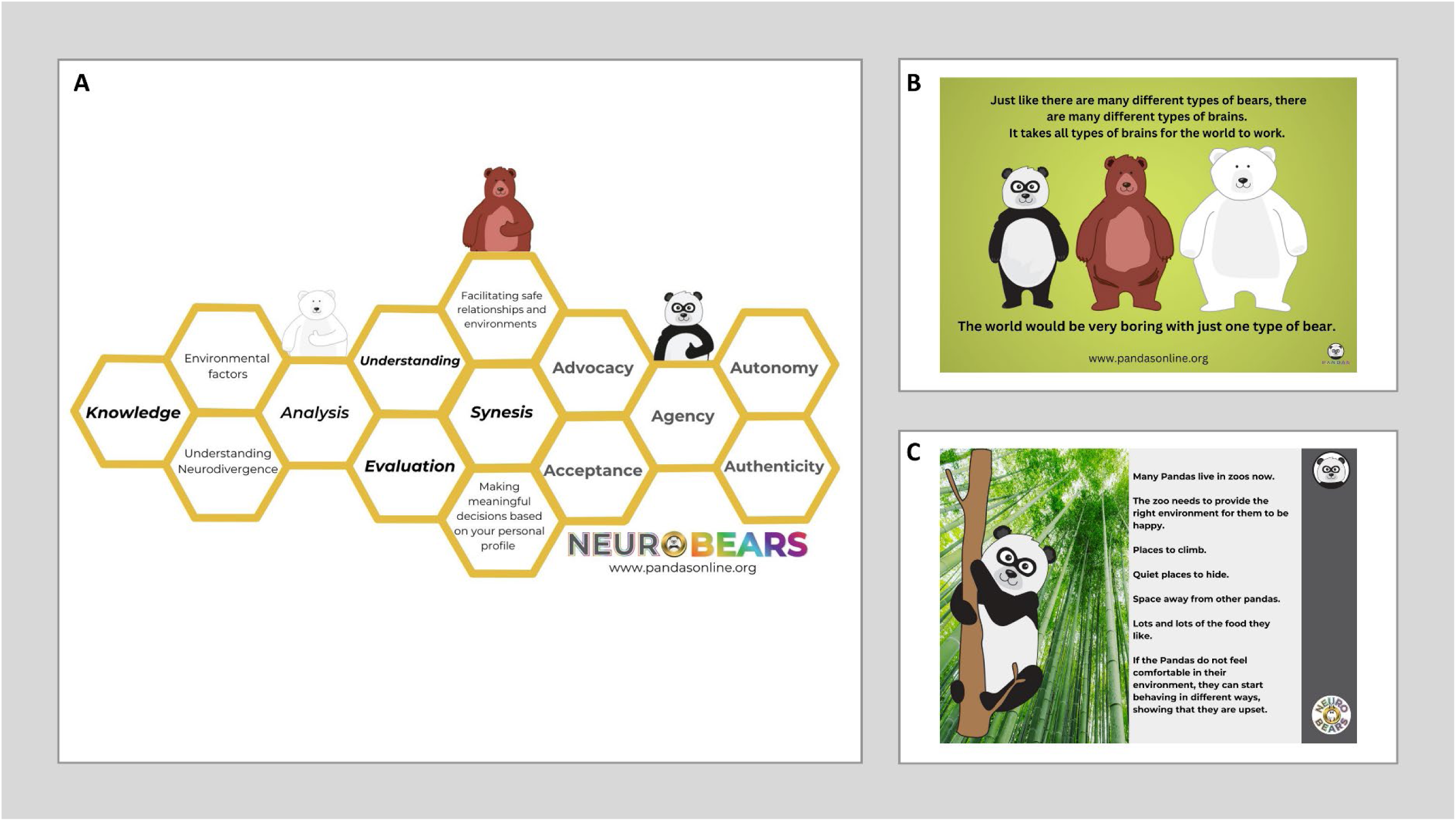
Information about the autism psychoeducation course NeuroBears (www.pandasonline.org), including topics covered (A) and use of bear analogies (amongst others) to support understanding of core concepts (B-C).

With compelling evidence and widespread calls for meaningful post-diagnostic support for autistic children and young people, this study aimed to address this significant gap in clinical practices and directly assess the benefit of an autistic-led psychoeducation autism course, developed within the social model of disability, and for autistic children (NeuroBears, https://www.pandasonline.org). We asked whether:

1. NeuroBears increased the children’s understanding of autism and their autistic experiences?
2. NeuroBears improved how children feel about being autistic?
3. NeuroBears facilitated children’s communication with others about their autistic experiences?
4. NeuroBears helped the children feel more able to and/or confident about self-advocating?
5. Was it important to children that NeuroBears was created and delivered by autistic individuals?
6. What did the children learn from NeuroBears?

## 2. Methods

### 2.1. Participants

We recruited children aged 8-14 years old (mean age=10.57, StDev=1.69; Figure 2A) and their parent/guardian. Parents had the option to nominate a safe adult committed to completing the psychoeducation course with the child in their home. For all participants, this was their parent.

**Figure 2.**
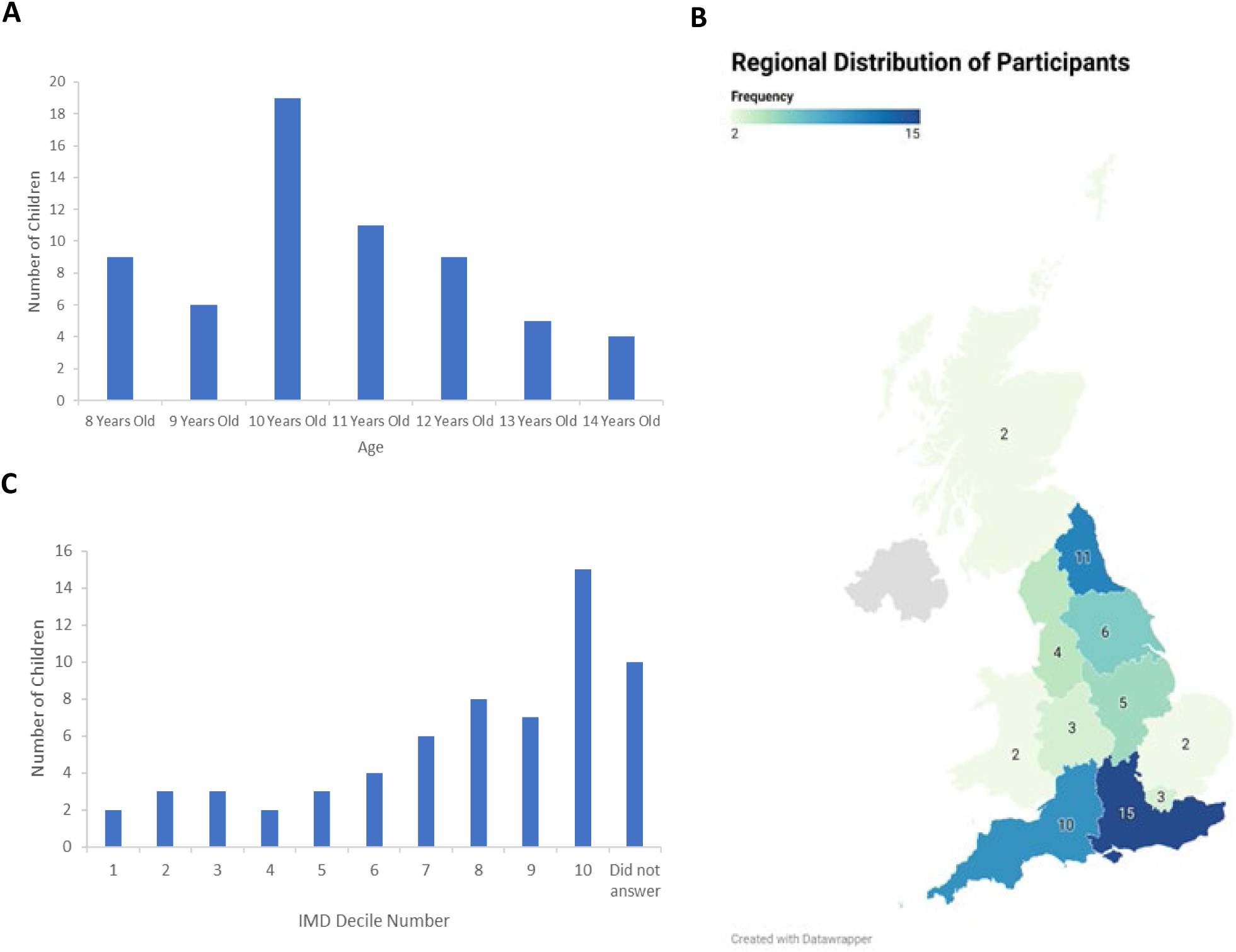
Flow chart of research and recruitment phases.

In total, 198 parents of autistic children initially expressed interest in participating in the study, resulting in 136 children consenting and completing the initial phase, and 72 children who completed both the pre- and post-NeuroBears questionnaires (see Figure 3 for full details). Finally, 9 of the 72 children’s data was excluded at this point, as they had completed less than 50% of the NeuroBears course. Hence, only the child data from the 63 children who completed both questionnaires and ≥50% of NeuroBears is reported here. 88.1% of this group completed 100% of the course.

**Figure 3.**
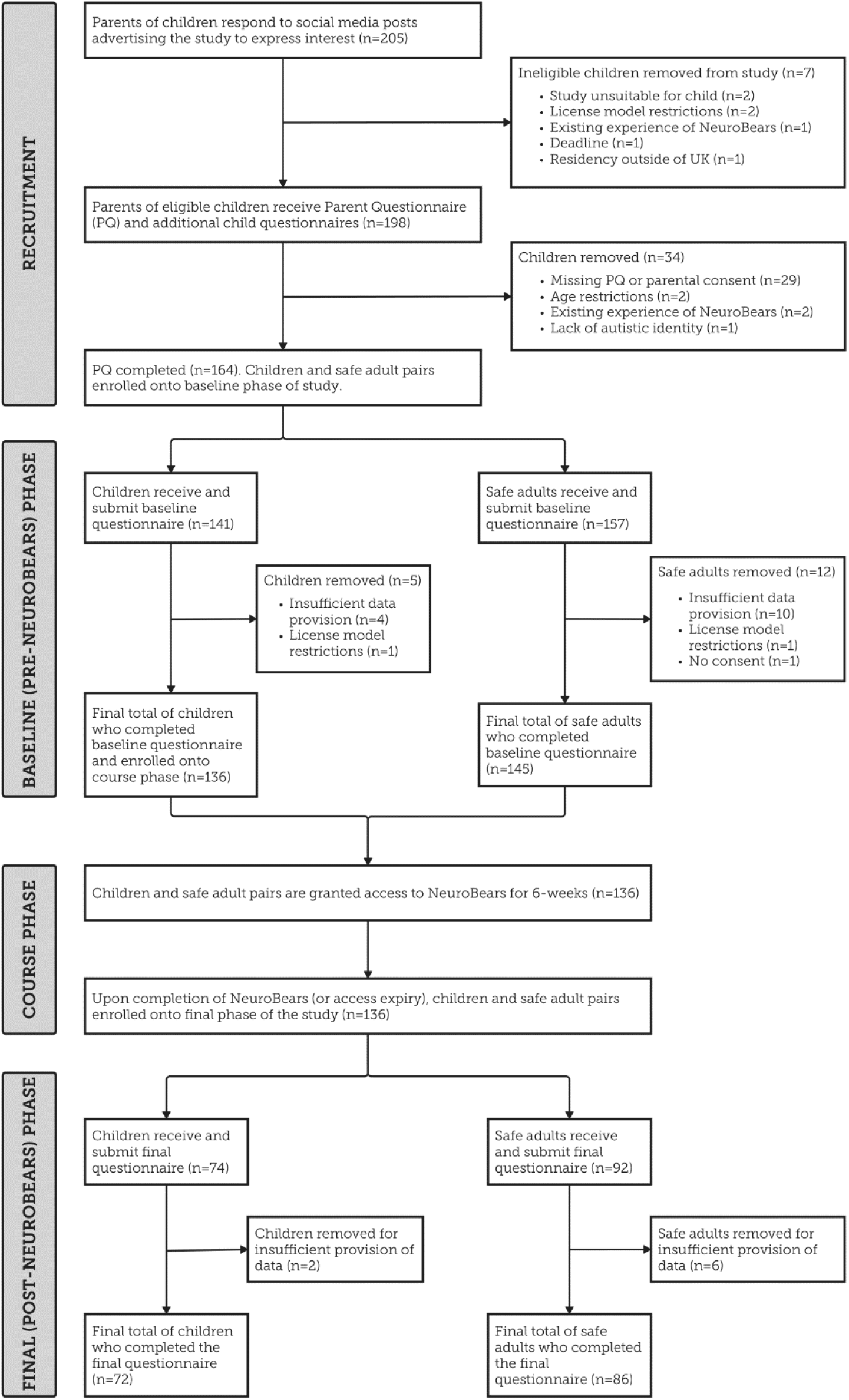
Participant Demographics. (A) Age distribution of the participants (n=63); (B) Areas of residence of participants ranked with respect to Index of Multiple Deprivation (IMD) decile scores. Decile 1 is the most deprived 10% of areas in England and decile 10 is the least deprived 10% of areas; (C) Regional distribution of participants.

Participants had a wide range of prior knowledge about autism prior to beginning the study (see Figure 4B). More specifically, pre-NeuroBears, 16 (of the 63) children endorsed the response options ‘I know a great deal’ or ‘I know a lot’ about their autistic brain. These 16 children were assigned to the ‘High Prior Knowledge’ group [mean age = 10.44 years (±1.50); sex assigned at birth = 62.5% female and 37.5% male]. Pre-NeuroBears, 33/63 children endorsed the ‘I know some things’ response option and were therefore assigned to the ‘Medium Prior Knowledge’ group [mean age = 10.75 years (±1.63); sex = 43.8% female and 56.3% male], whilst 14/63 children endorsed the ‘I know a little’ or ‘I know nothing’ options at baseline and were assigned to the ‘Low Prior Knowledge’ group [mean age = 10.5 years (±2.03); sex = 42.9% female and 57.1% male]. There was no significant age difference between these 3 groups [F (2,59) = 0.221, *p* = .802].

**Figure 4.**
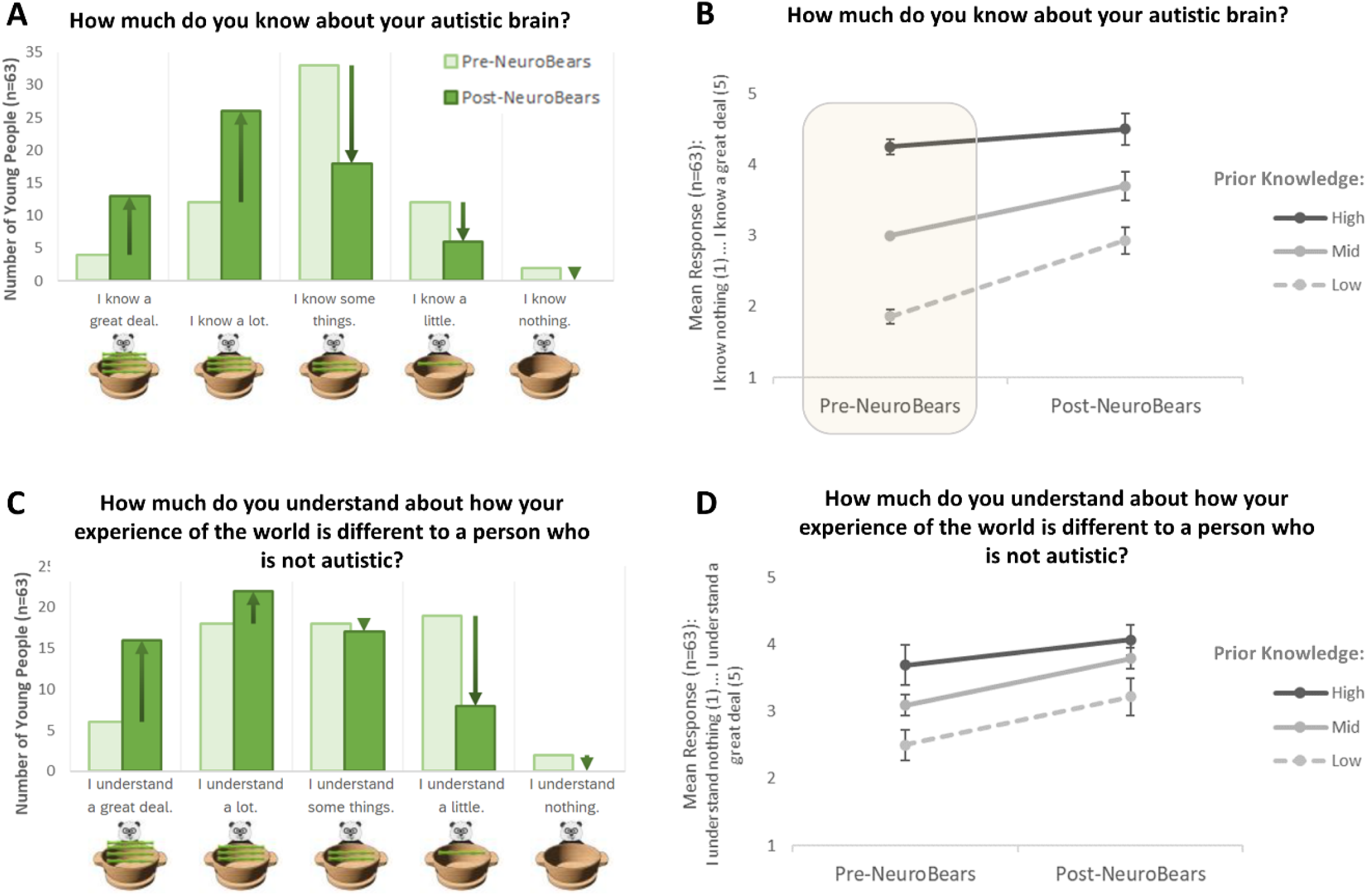
Children’s understanding of autism (I): (A-B): “How much do you know about your autistic brain?”; (C-D): “How much do you understand about how your experience of the world is different to a person who is not autistic?”. A and C represent the pre- and post-NeuroBears responses at the level of the individual response options, whilst B and D show the overall change in responses provided by the children pre- and post-NeuroBears with respect to group (i.e., High, Medium, and Low prior knowledge about autism). Note that overlaid box is in Panel B is to emphasise the need for caution when interpreting between-group and interaction effects on this parameter, as the pre-NeuroBears responses for this specific question were used to differentiate the three groups in the first instance.

Participants were all residing in the UK (regional distribution shown in Figure 2B). 93.7% of the children’s parents were their biological parents, 3.2% were their adoptive parents, and 1.6% were their foster parents. 47.6% of the children identified their gender as boy, 49.2% as girl, 1.6% as non-binary, and 1.6% self-described. 92.1% of participants were white, and 7.9% of participants were of mixed or multiple ethnic groups. The socioeconomic status (SES) of children was interpreted through using the Index of Multiple Deprivation (IMD) (28) i.e., area-based deprivation indices (based on a combination of seven different types of deprivation) which rank the population from most deprived to least deprived. A mean IMD decile (1=most deprived… 10=least deprived) was calculated as 7.23 (SD = 2.76) (i.e., less deprived than the UK average). An overview of the IMD distribution is shown in Figure 2C. As per inclusion criteria, all children were autistic, established via formal diagnosis or self-identification. The majority had at least one co-occurring condition (see Table 1). The average time since autism diagnosis was 2.26 years (StDev=2.42).

**Table 1.**
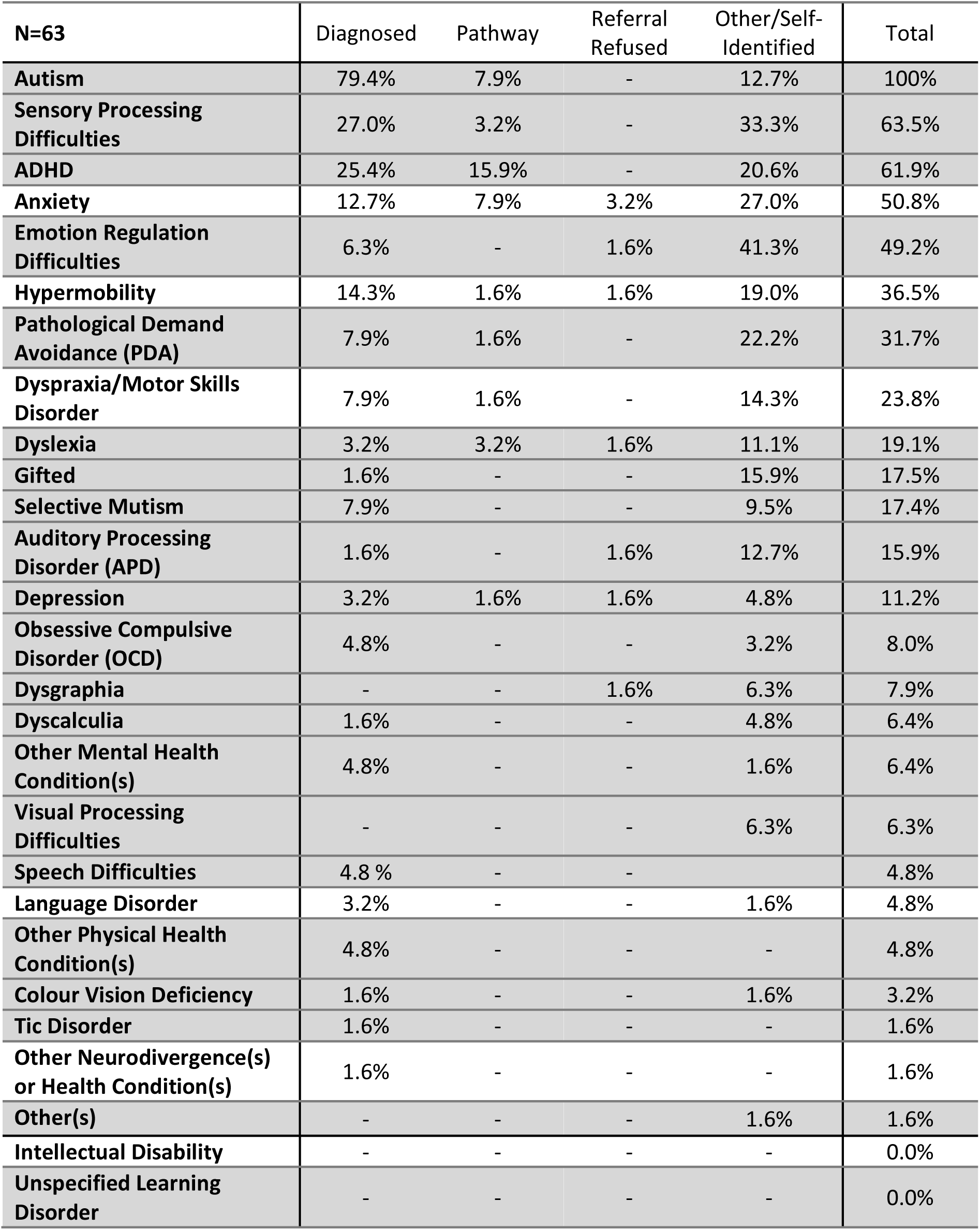
Percentages of the 63 children who completed both questionnaires (pre- and post-NeuroBears) and at least 50% of NeuroBears course itself.

### 2.2. Research Ethics and Language

This study was approved by the Faculty of Medical Sciences Research Ethics Committee, part of Newcastle University’s Research Ethics Committee. Consistent with endorsements from autistic adults in the United Kingdom (Kenny et al., 2016), we use identity-first language within this paper (e.g., ‘autistic child’ rather than ‘child with autism’).

### 2.3. Design

The study used a concurrent embedded mixed-methods within-participants design, within which qualitative data was collected to supplement quantitative data. Both the children and their Safe Adults completed the pre-NeuroBears questionnaire prior to receiving access to the NeuroBears course materials, and the post-NeuroBears questionnaire approximately 6 weeks later.

A participatory approach was taken throughout the research. This involved frequent meetings with NK (creator and co-author of NeuroBears) throughout the design and write-up phases for advice with respect to wording of questions, language use, accessibility, etc.

### 2.4. Materials

Questionnaires were developed by the researchers at Newcastle University (SM, SC and HC, with advice from JR). NK also provided significant input into the wording of questions [e.g., with respect to the accessibility of the questions for the target age group (8–14-year-olds) and whether this aligned with the language used in NeuroBears] and whether the questions developed fully aligned with, and encompassed, the learning objectives of NeuroBears.

The parent/guardian questionnaire gathered information with respect to the children, including age, sex, gender, ethnicity, age at time of autism diagnosis, other neurodivergent and health conditions, and family demographics, including parent/sibling diagnoses of autism. Data from the Safe Adult questionnaires will be reported elsewhere but roughly mirrored the children’s questionnaires.

The baseline (pre-NeuroBears) questionnaires focused on the children’s understanding and feelings about being autistic. It also asked children to reflect on how they communicate with others with respect to their autistic experiences, such as whether they do talk to others about being autistic and how they feel when they do this and how brave they feel to self-advocate. These same questions were repeated in the post-NeuroBears questionnaire, after which the children were also asked to describe how they found completing NeuroBears and specific questions about various aspects of its content. As a unique aspect of NeuroBears as a psychoeducation course for autistic children is that it was created, written, and delivered by autistic people, one of these specific questions asked whether this was important to the children, and a second focused on what they learnt during NeuroBears. Free-text response boxes were available throughout both the pre- and post-NeuroBears questionnaires, so that the children were able to expand on their responses. For the full list of questions reported here, see Supplemental Materials (pages 2-4).

### 2.5. NeuroBears Course

NeuroBears is a pre-recorded online course, with accompanying guides and workbooks, for autistic children, and is all about the autistic experience. It was created by autistic adults in collaboration with autistic young people and is not affiliated with any health or academic organisation. It provides psychoeducation to autistic children aged 8-14 years, alongside their nominated safe-adult, and covers the topics described in (Figure 1A), using bear analogies (amongst others) to support understanding of core concepts (Figure 1B-C). NeuroBears was created with a vision of neutrality, a vision of creating lifelong learning and inspiring curiosity in autistic young people about themselves and their neurodivergence. It seeks to recognise that there are many strengths and struggles to being autistic, and supports children to find and recognise these in themselves, within the many deficit-based systems and various intersectionalities that they live in.

### 2.6. Procedure

Participants volunteered by responding to recruitment posts on the PANDAS Online social media pages and the Newcastle University Cognitive Developmental Lab’s Facebook page. Once parents expressed interest in the research, they were emailed the initial Parent questionnaire (by AW and CE who were responsible for data collection). Sibling questionnaires were also provided if applicable.

Upon receipt of initial parental consent, the pre-NeuroBears child and Safe Adult questionnaires were emailed to the safe adult. Once the Initial Parent, Safe-Adult, and Child questionnaires were successfully completed, virtual access to the NeuroBears course was provided via the website https://www.pandasonline.org/. Participants then had 43 days of online access. After they had informed the research team that they had completed the course, or their access had expired, the links to the post-NeuroBears questionnaires was provided. Reminders were emailed roughly weekly (for around 6 weeks maximum) to prompt participants to complete the questionnaires, and the NeuroBears course.

To support participation, children were encouraged to complete both the pre- and post-NeuroBears questionnaires via a video call with one of the researchers. The researcher worked through the questionnaire alongside the young person, inputting their responses. If they did not wish to complete the questionnaire with a researcher, they completed it in their own time.

After data had been collected and analysed, participants (both children and adults) were invited to attend a presentation of the early findings.

### 2.7. Data Analysis

Quantitative analysis was conducted using IBM SPSS Statistics (version 29.0.1.0). All response options (apart from the two questions probing specific emotions) used a 5-point Likert scale (see Supplemental Information). A value of 1 was given to the most negative response option and a value of 5 was given to the most positive response option. Each core question was completed both before and after NeuroBears.

Mixed factorial 3 x 2 ANOVAs (Group=3 levels: Low, Medium or High Prior Knowledge) x Timepoint=2 levels: Pre-NeuroBears or Post-NeuroBears) were implemented through the repeated measures general linear model for all core Likert scale questions answered pre- and post-NeuroBears (29, 30).

Some questions related to specific emotions experienced and required a dichotomous (Yes/No) response option. Comparison of these responses, pre- and post-NeuroBears, were analysed using the McNemar test. In addition, and for these specific questions, the total number of emotions related to positive affectivity (i.e., Happy, Comfortable, Calm, Excited, Accepting, Engaged and Okay) and negative affectivity (i.e., Numb, Tired, Sad, Angry, Frustrated, Scared, Anxious, Depressed, Unwell, Panicked and Nervous) were summed for each participant. Mixed factorial 3 x 2 x 2 ANOVAs Group (3 levels: Low, Medium or High Prior Knowledge) x Timepoint (2 levels: Pre-NeuroBears or Post-NeuroBears) x Valence (2 levels: Positive Affectivity and Negative Affectivity) were implemented through the repeated measures general linear model. If indicated, paired-samples t-tests used to compare the pre- and post-NeuroBears sum totals for positive and negative affectivity separately.

Mixed factorial 3 x 2 x 3 ANOVAs Group (3 levels: Low, Medium or High Prior Knowledge) x Timepoint (2 levels: Pre-NeuroBears or Post-NeuroBears) x Context (3 levels: People at Home, Friends/Peers, Teachers) were implemented through the repeated measures general linear model. If indicated, related-samples Friedman’s analysis of variance by ranks with Dunn-Bonferroni post hoc tests were employed to explore differences across contexts (i.e., at home, when out and about, and at school) at specific timepoints. Where free text responses provide wider context and further understanding of data, these are included in the Discussion section. A thematic analysis of the free text responses will be published elsewhere.

## 3. Results

### 3.1. Understanding of Autism

Significant main effects of NeuroBears were found for all items relating to the children’s understanding of autism and their autistic experiences: “How much do you know about your autistic brain?” [F(1, 60) = 42.649, *p* < .001] (Figure 4A-B), “How much do you understand about how your experience of the world is different to a person who is not autistic?” [F(1, 60) = 12.196, *p* < .001] (Figure 4C-D), “Do you think that you have any co-occurring/other conditions, as well as being autistic?” [F(1, 60) = 6.820, *p* = .011], “Do you think there are any good things about being autistic?” [F(1, 59) = 2.920, *p* = .017] (Figure 5A-B), “Do you feel you are good at some things because you are autistic?” [F(1, 59) = 5.979, *p* = .017] (Figure 5C-D), and “Do you feel some things are harder for you because you are autistic?” [F(1, 60) = 22.238, *p* < .001] (Figure 5E-F). All changes were in the positive direction (see Figures 2 and 3).

**Figure 5.**
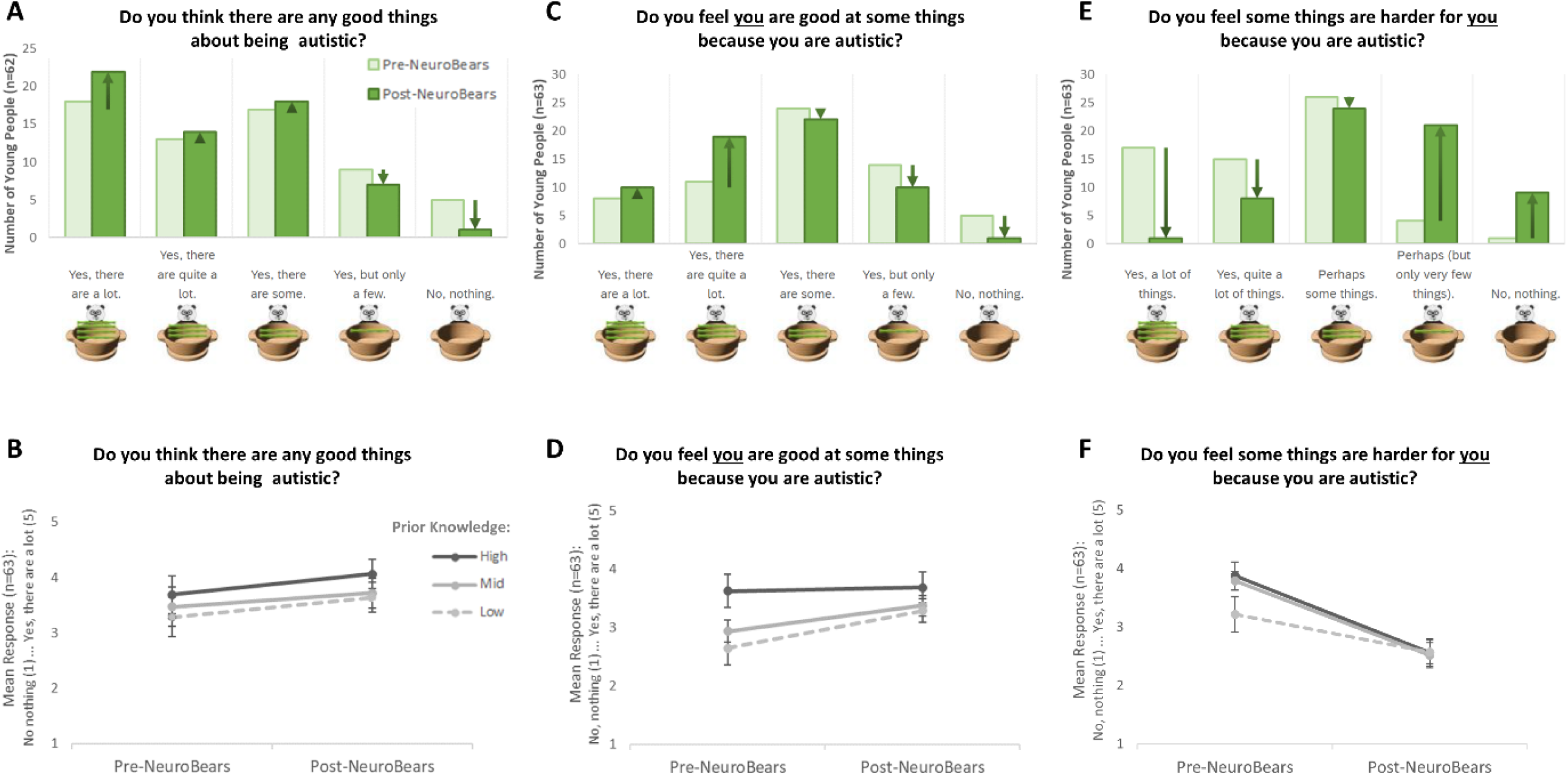
Children’s understanding of autism (II): (A-B): “Do you think there are any good things about being autistic?”; (C-D): “Do you feel you are good at some things because you are autistic?”; (E-F): “Do you feel some things are harder for you because you are autistic?”. Panels A, C and E represent the pre- and post-NeuroBears responses at the level of the individual response options, whilst panels B, D and F show the overall change in responses provided by the children pre- and post-NeuroBears with respect to group.

A significant main effect of Group was found for the item “How much do you know about your autistic brain?” [F(2, 60) = 4.386, *p* = .017], alongside a significant Timepoint x Group interaction [F(2, 60) = 92.356, *p* < .001] (see Figure 4B). A significant main effect of Group was also found for the item “How much do you understand about how your experience of the world is different to a person who is not autistic?” [F(2, 60) = 7.198, *p* = .002], but no significant Timepoint x Group interaction. No other significant between-group or significant Timepoint x Group interactions were observed.

### 3.2. Communicating about Being Autistic

A significant main effect of NeuroBears was found for the item “Do you feel you have the words to talk about being autistic?”; F(1, 60) = 22.647, *p* < .001). A significant main effect of Group [F(2, 60) = 6.742, *p* = .002] and a significant interaction between Group x NeuroBears F(2, 60) = 3.834, *p* = .027] were also found (Figure 6B).

**Figure 6.**
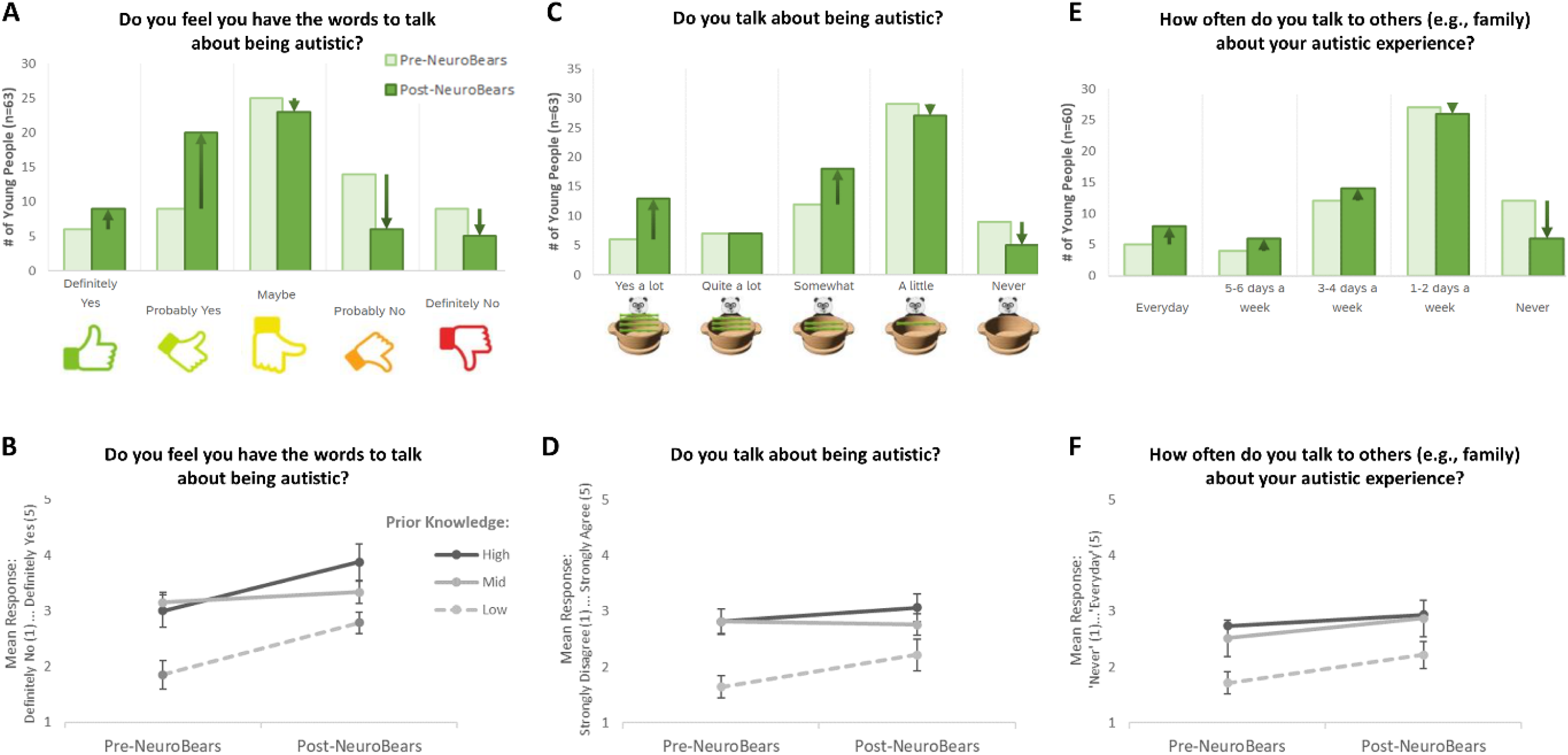
Communicating about being autistic (I): (A-B): “Do you feel you have the words to talk about being autistic?”; (C-D): “Do you talk about being autistic?”; (E-F): “How often do you talk to others (e.g., family) about your autistic experience?”. Panels A, C and E represent the pre- and post-NeuroBears responses at the level of the individual response options, whilst panels B, D and F show the overall change in responses provided by the children pre- and post-NeuroBears with respect to group.

When asked directly “Do you talk about being autistic”, there was a no significant main effect of NeuroBears [F(1, 60) = 3.627, *p* = .062] or statistically significant interaction between Group x NeuroBears [F(2, 60) = 2.112, *p* = .130]. A significant main effect of Group was found [F(2, 60) = 5.183, *p* = .008], driven by the group with the lowest prior autism knowledge talking least about being autistic (see Figure 6C-D). When asked “How often do you talk to others (e.g., family) about your autistic experience?” (Figure 6E), there was a significant main effect of NeuroBears [F(1, 57) = 7.3017, *p* = .009] (Figure 6F). No main effect of Group [F(2, 57) = 3.134, *p* = .051] or Group x NeuroBears interaction [F(2, 57) = 0.365, *p* = .696] were evident.

When considering the question “How comfortable do you feel talking about being autistic with your family ”, a significant main effect of NeuroBears [F(1, 57) = 14.889, *p* < .001], alongside a significant main effect of Group [F(2, 57) = 7.867, *p* < .001] and a significant Group x NeuroBears interaction [F(2, 57) = 3.203, *p* = .048], were found (Figure 7A-B). This pattern was not repeated for the questions “How comfortable do you feel talking about being autistic with other people (excluding family)?” – with no significant main effect of NeuroBears [F(1, 54) = 2.807, *p* = .100] or significant Group x NeuroBears interaction [F(2, 54) = 0.234, *p* = .792]. A significant main effect of Group [F(2, 54) = 9.000, *p* < .001] indicated that those with the least knowledge of autism entering the research, were the most uncomfortable when talking about being autistic with people outside of their family throughout the research (Figure 7C-D).

**Figure 7.**
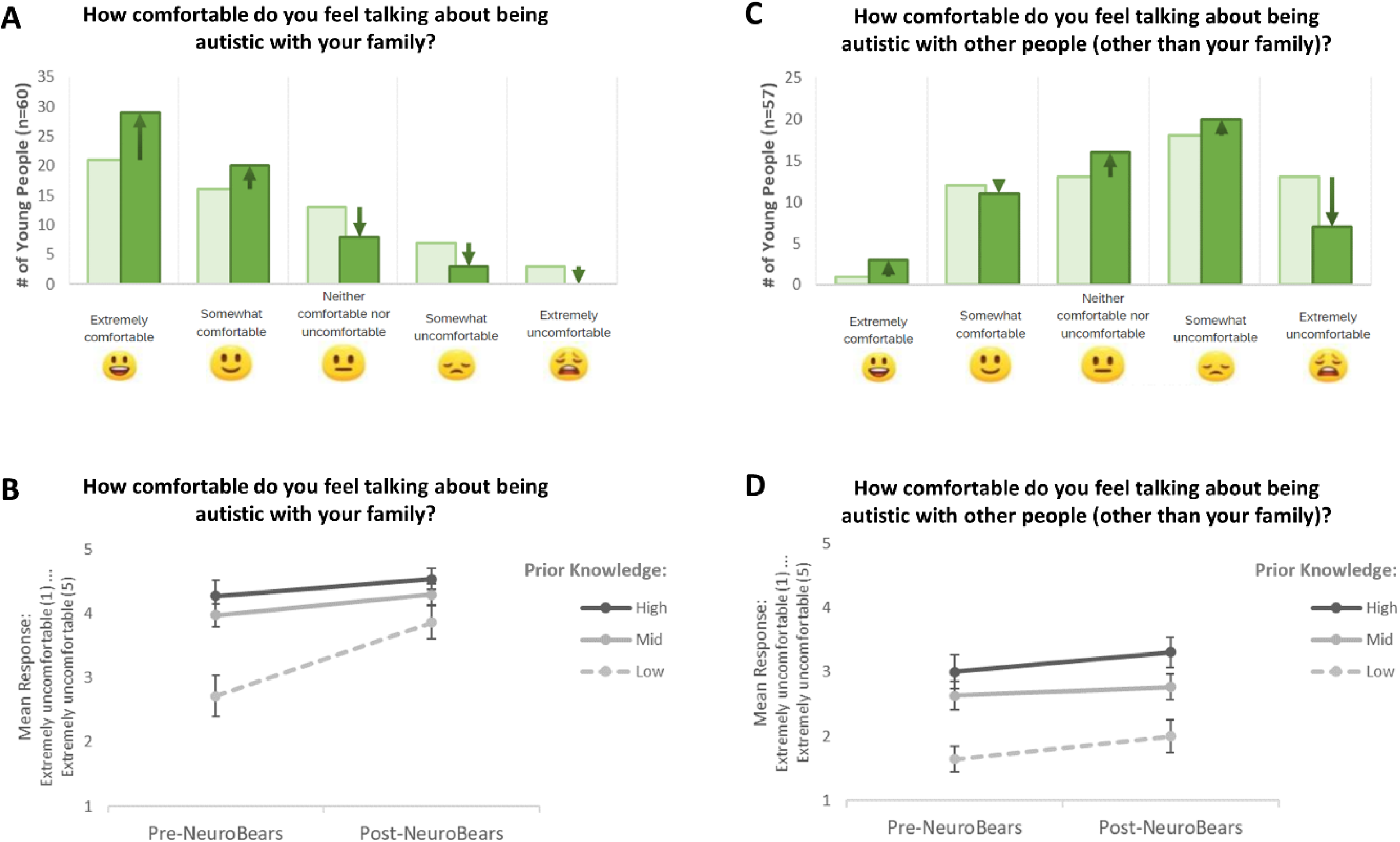
Communicating about being autistic (II): (A-B): “How comfortable do you feel talking about being autistic with your family?”; (C-D): “How comfortable do you feel talking about being autistic with other people (other than your family)?”. Panels A and C represent the pre- and post-NeuroBears responses at the level of the individual response options, whilst panels B and D show the overall change in responses provided by the children pre- and post-NeuroBears with respect to group.

We also explored the specific emotions that autistic children experience when talking to other people about being autistic. Both pre- and post-NeuroBears, the emotion most frequently endorsed by participants was ‘Anxious’ (Figure 8A). When each of the 18 emotions listed were considered individually, no statistically significant differences in the frequency of any single emotion endorsed by participants pre-relative to post-NeuroBears were found.

**Figure 8.**
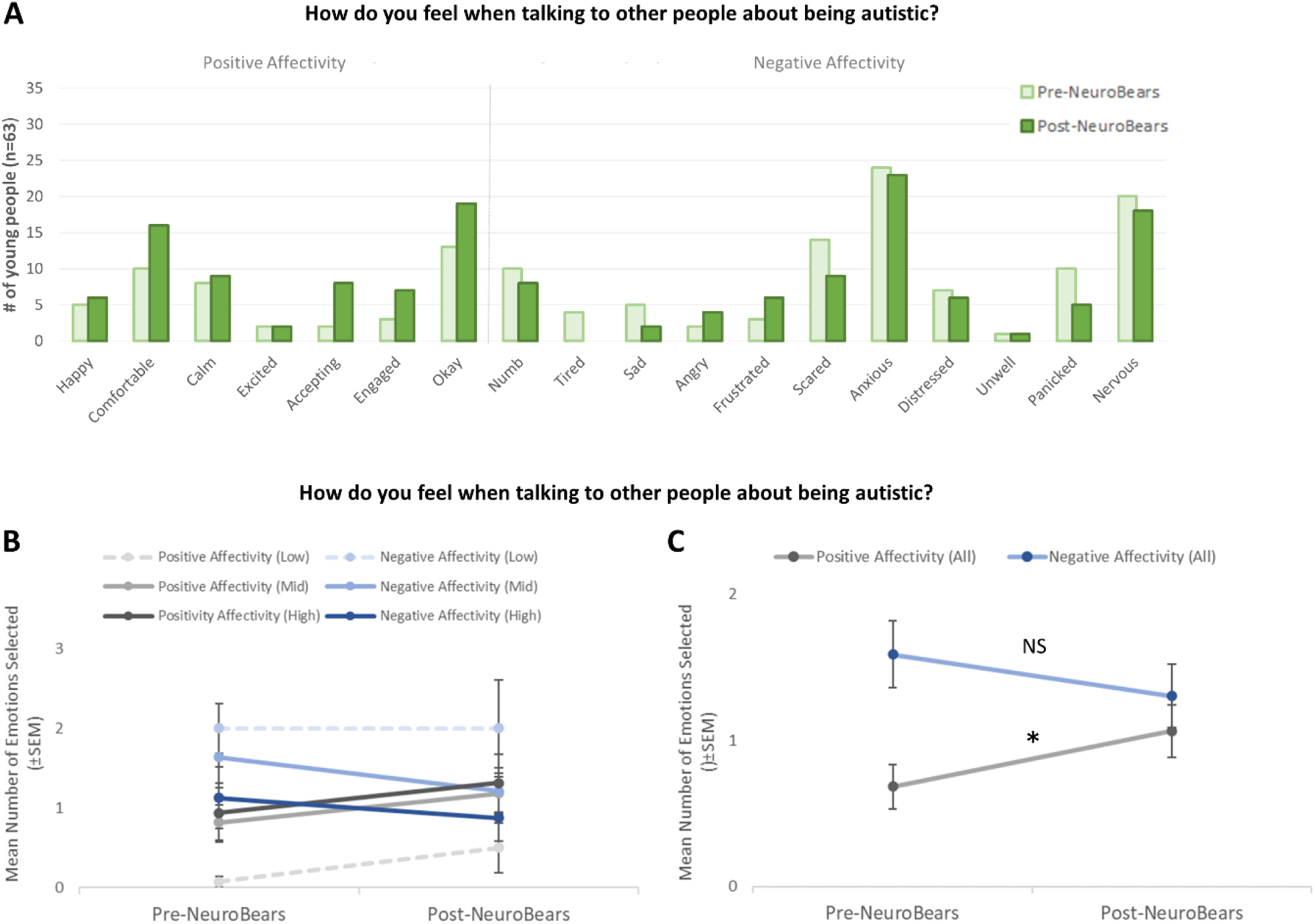
Feelings experienced when communicating about being autistic (I): (A) Emotions endorsed by the children when asked how they feel when talking to other people about being autistic pre- and post-NeuroBears; (B) Summed total of endorsed emotions relating to positive and negative affectivity pre- and post-NeuroBears broken down with respect to Prior Knowledge of Autism Group factor; (C) Overall summed total of endorsed emotions relating to positive and negative affectivity pre- and post-NeuroBears. * p < .05; NS = non-significant change.

When all endorsed emotions were summed, a significant overall effect of NeuroBears [F(1, 60) = 9.766, *p* = .002], and a significant Group x NeuroBears interaction [F(2,60) = 5.293, *p* = .008], were found. Hence, the number of emotions identified changed between the pre- and post-NeuroBears questionnaires, and this change was different across the three groups (Low, Medium and High Prior Knowledge; Figure 8B). No Group x NeuroBears x Emotion Valence interaction was observed. When collapsed across group, a paired samples t-test found that overall, more emotions associated with positive affectivity were endorsed by the children post-NeuroBears relative to pre-NeuroBears [t(62) = -1.965, *p* < .05; Figure 8C].

Despite the above significant change in positive affect, there was no significant main effect of NeuroBears when children were asked to report how brave they feel talking about being autistic [F(1, 60) = 2.325, *p* = .133] (Figure 9A-B). However, when this was re-framed to think about safety instead of feelings of bravery and referred explicitly to discussions with family members (Figure 9C), there was a significant main effect of NeuroBears [F(1, 56) = 5.505, *p* = .023], indicating that the children reported feeling safer talking to others such as family post-NeuroBears relative to pre-NeuroBears. Interestingly, for both questions, there was a significant main effect of Group [F(2, 60)= 3.276, *p* = .045 and F(2, 56)= 6.712, *p* = .002 respectively], with the children with the least knowledge of autism prior to the research feeling less brave and less safe across the two timepoints (Figure 9B and 9D). No significant Group x NeuroBears interactions were observed for either item.

**Figure 9.**
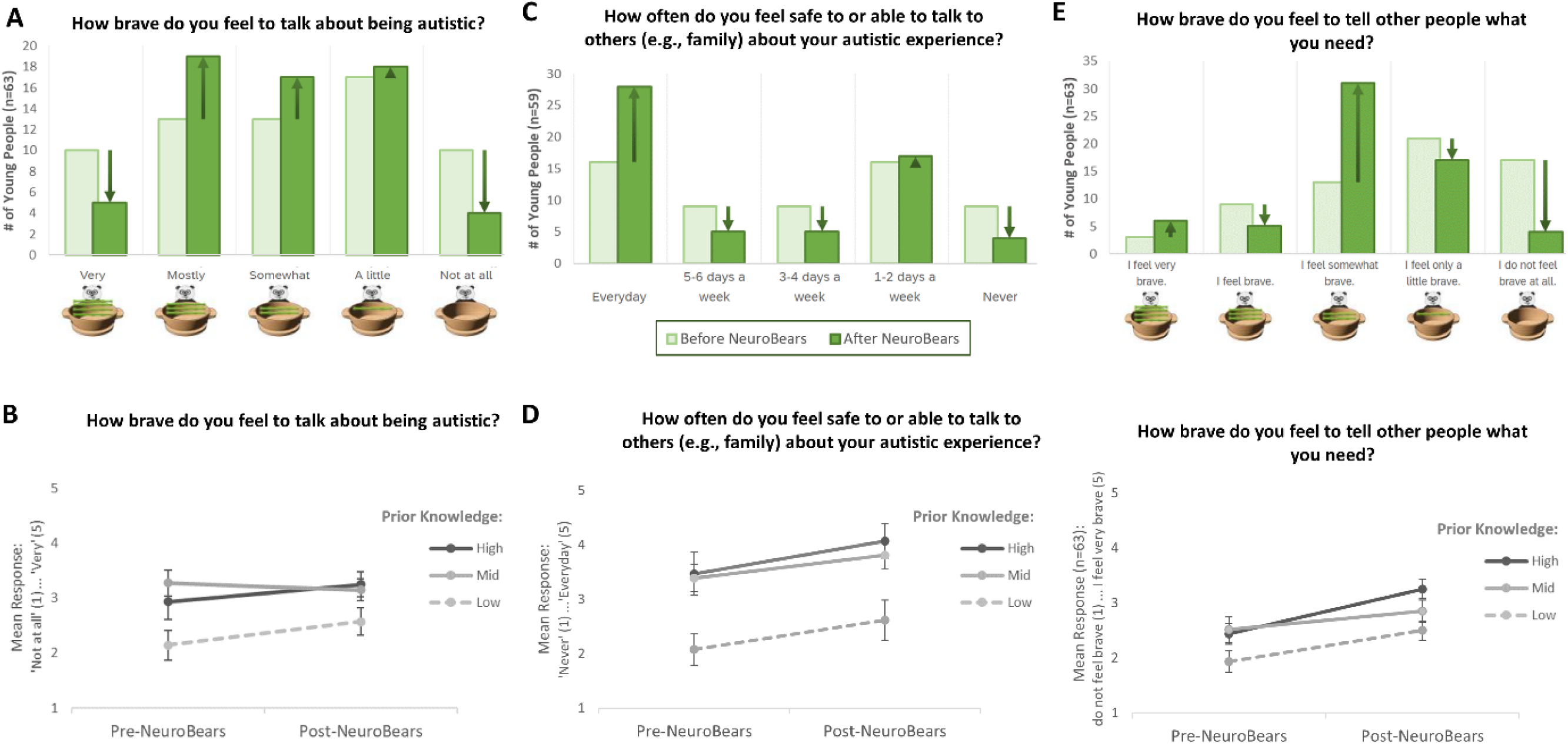
Feelings experienced when communicating about being autistic (II). (A-B): “How brave do you feel to talk about being autistic?”; (C-D): “How often do you feel safe to or able to talk to others (e.g., family) about your autistic experience?”; (E-F): “How brave do you feel to tell other people what you need?”. Panels A, C and E represent the pre- and post-NeuroBears responses at the level of the individual response options, whilst panels B, D and F show the overall change in responses provided by the children pre- and post-NeuroBears with respect to group.

### 3.3. Self-Advocacy

Significant main effects of NeuroBears were found for both items relating to self-advocacy: “How brave do you feel to tell other people what you need?” [F(1, 60) = 15.132, *p* < .001] (Figure 9E-F) and “Do you always feel brave enough to tell others what you need?” [F(1, 59) = 7.625, *p* = .008]; with 41.9% of children reporting feeling brave enough to tell people what they need ‘Most of the time’ or ‘All of the time’ post-NeuroBears, relative to only 27% of children pre-NeuroBears.

### 3.4. Feelings about Being Autistic

A significant main effect of NeuroBears was found for the item “I like being autistic” [F(1, 60) = 7.340, *p* = .009], coupled with no significant main effect of Group [F(2, 60)=1.760, p = .181] or Group x NeuroBears interaction F(2, 60)=0.598, *p* = .553]. Hence, regardless of Group, the children endorsed the statement that they like being autistic more strongly post-NeuroBears relative to pre-NeuroBears (Figure 10A-B).

**Figure 10.**
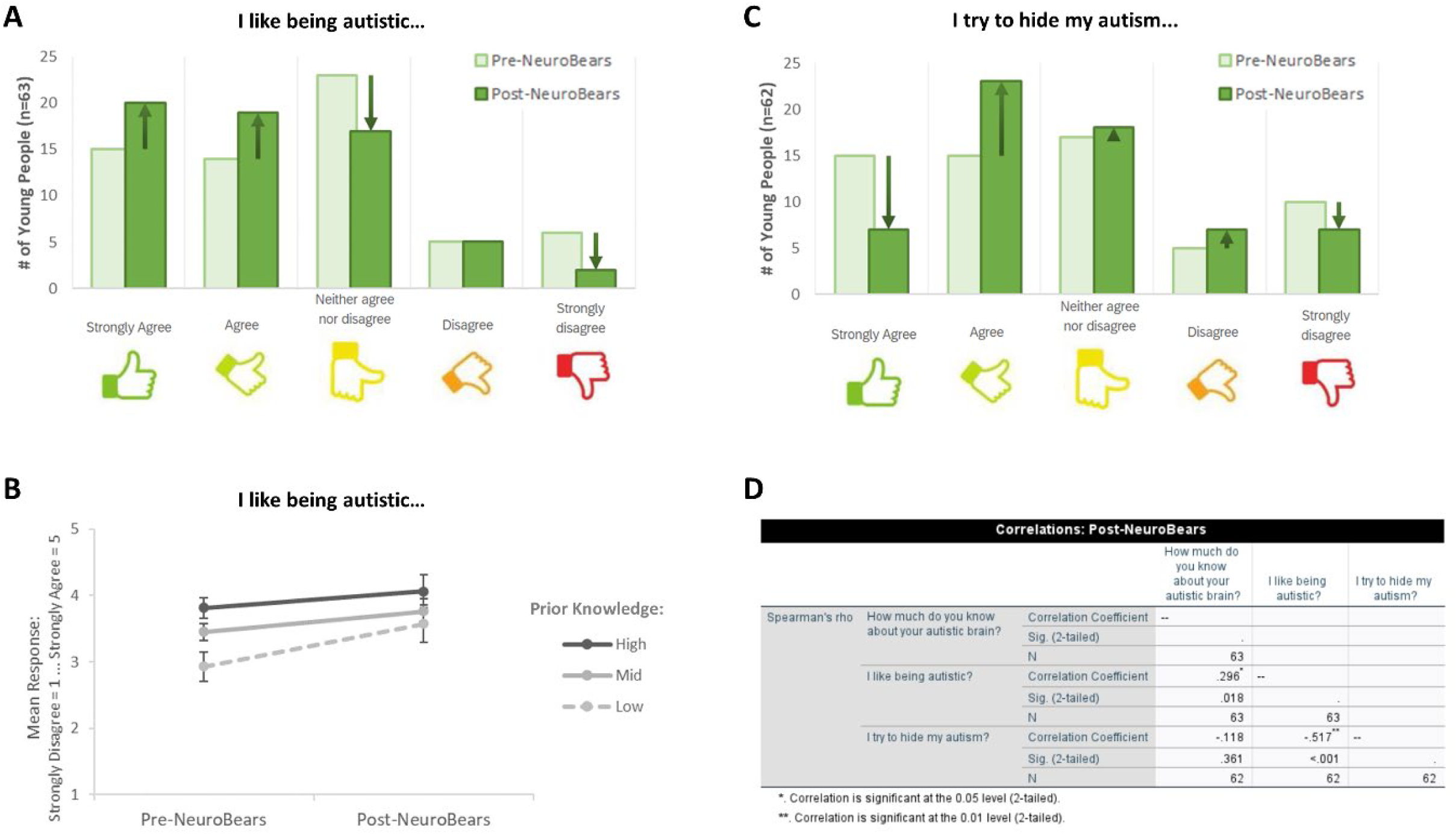
Autistic Identity and Masking. (A-B): “I like being autistic?”; (C): “I try to hide my autism?”. Panels A and C represent the pre- and post-NeuroBears responses at the level of the individual response options, whilst panel B shows the overall change in responses provided by the children pre- and post-NeuroBears with respect to group. (D) Correlation matrix exploring the relationship between the children’s understanding of being autistic, their feelings about being autistic, and how much they mask posts-NeuroBears.

No significant main effect of NeuroBears was found for the item “I try to hide my autism” [F(1, 59) = 0.105, *p* = .747], nor a significant main effect of Group [F(2, 59) = 0.329, *p* = .721] or Group x NeuroBears interaction [F(2, 59) = 0.029, *p* = .972] (Figure 10C).

Spearman’s rho correlations were conducted to evaluate the relationship between the children’s knowledge of their autistic brains, how much they like being autistic, and how much they try to hide their autism both pre- and post-NeuroBears. A significant positive relationship between how much the children know about their autistic brains and how much they like being autistic (Pre-NeuroBears: r_s_ = 0.257, *p* = .042; Post-NeuroBears: r_s_ = 0.294, *p* = .018), and a significant negative relationship between how much the children like being autistic and how much they try to hide their autism (Pre-NeuroBears: r_s_ = -0.364, *p* = .003; Post-NeuroBears: r_s_ = -0.517, *p* < .001) (Figure 10D and Figure S1), were found.

Exploring in more detail how participants feel about being autistic, the most frequently endorsed emotion selected by the children, both pre- and post-NeuroBears, was ‘Okay’ (Figure 11A), followed by ‘Comfortable’ (2^nd^), ‘Accepting’ (3^rd^) and ‘Happy’ (4^th^). The 5^th^ most frequently endorsed emotion pre-NeuroBears was ‘Frustrated’, but this was replaced with ‘Calm’ post-NeuroBears. ‘Numb’ was the one emotion which showed a significant change, moving from 6^th^ position (pre-NeuroBears) to 12^th^ position (Post-NeuroBears) (*p* = .013; Figure 11A). No other significant differences in individual emotions were observed, although ‘angry’ showed a trend towards significance (*p* = .063).

**Figure 11.**
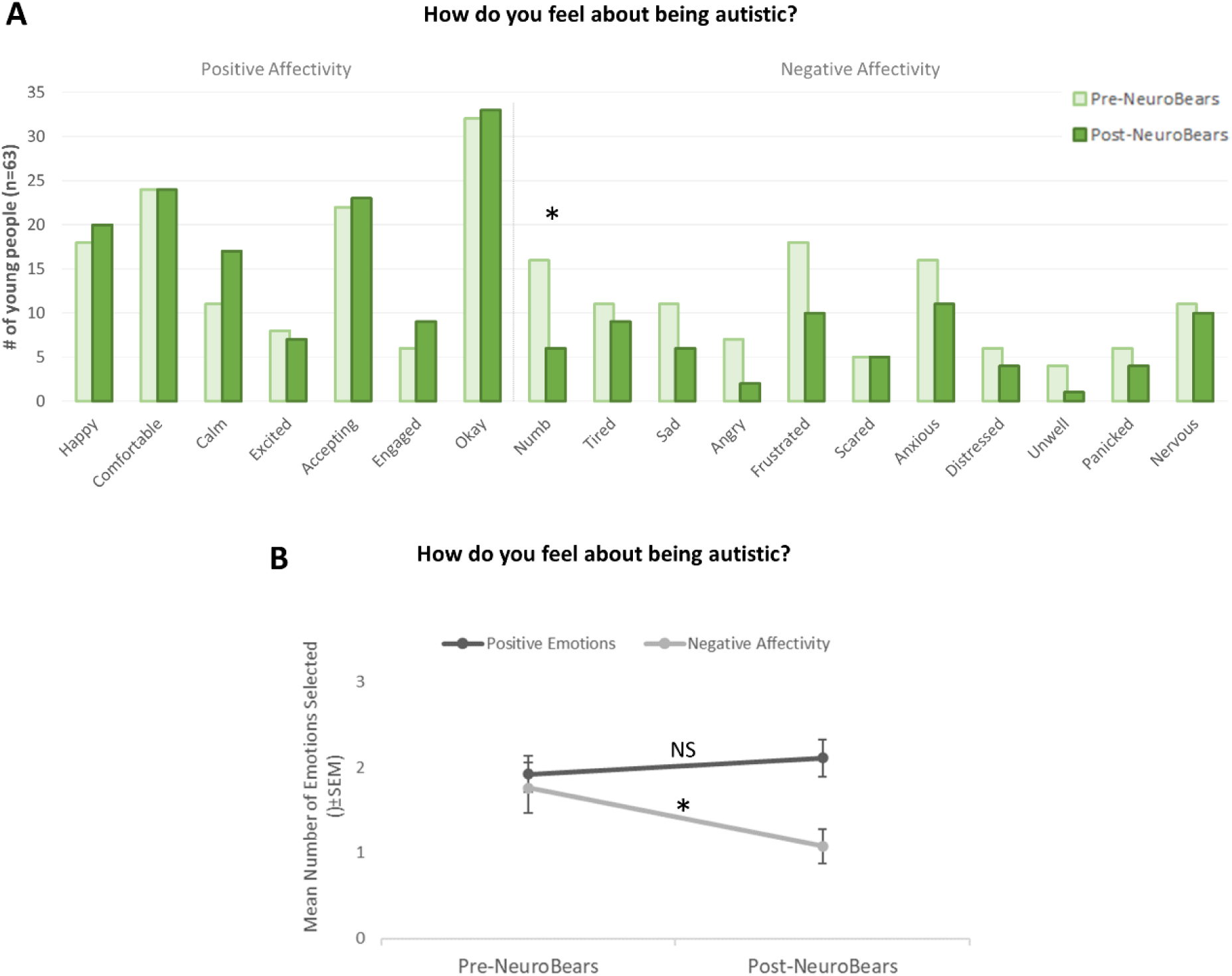
Autistic Identity (II). (A) Emotions endorsed by the children when asked how they feel about being autistic pre- and post-NeuroBears; (B) Overall summed total of endorsed emotions relating to positive and negative affectivity pre- and post-NeuroBears. * *p* < .05; NS = non-significant change.

When all endorsed emotions were summed, the results of the 3 x 2 x 2 mixed factorial ANOVA found no significant overall effect of NeuroBears [F(1, 60) = 0.243, *p* = .624]. However, there was a significant main effect of Emotion Valence [F(1, 60) = 4.856, *p* = .031] and a significant NeuroBears x Emotion Valence interaction [F(1,60) = 4.045, *p* = .049]. Hence, the relative number of emotions associated with positive affect relative to the number of emotions associated with negative effect varied significantly post-NeuroBears relative to pre-NeuroBears, with the balance between positive and negative emotions similar pre-NeuroBears but different post-NeuroBears (see Figure 11B); with paired samples t-tests revealing a statistically significant reduction in negative affect post-NeuroBears relative to pre-NeuroBears (t(62) = 2.335, *p* < .05).

<u>Impact of Context (Home, Friends, School)</u>: We also explored how the children’s feelings about being autistic may change in response to who they are with. More specifically, we focused on how the children feel about being autistic when they are with ‘People at Home’, ‘Friends/Peers’, and ‘People at School’. Mirroring the findings from phase 1 (n=136; see Mullally et al., in preparation) in the current subgroup of children who completed all phases (n=63), the results of the 3 (Group) x 2 (NeuroBears) x 3 (Context) mixed factorial ANOVA found no significant overall main effects of NeuroBears [F(1, 59) = 0.717, *p* = .401] or Group [F(2, 59) = 2.764, *p* = .071] (Figure 12A-D). However, there was a significant main effect of Context [F(2, 60) = 39.491, *p* < .001].

**Figure 12.**
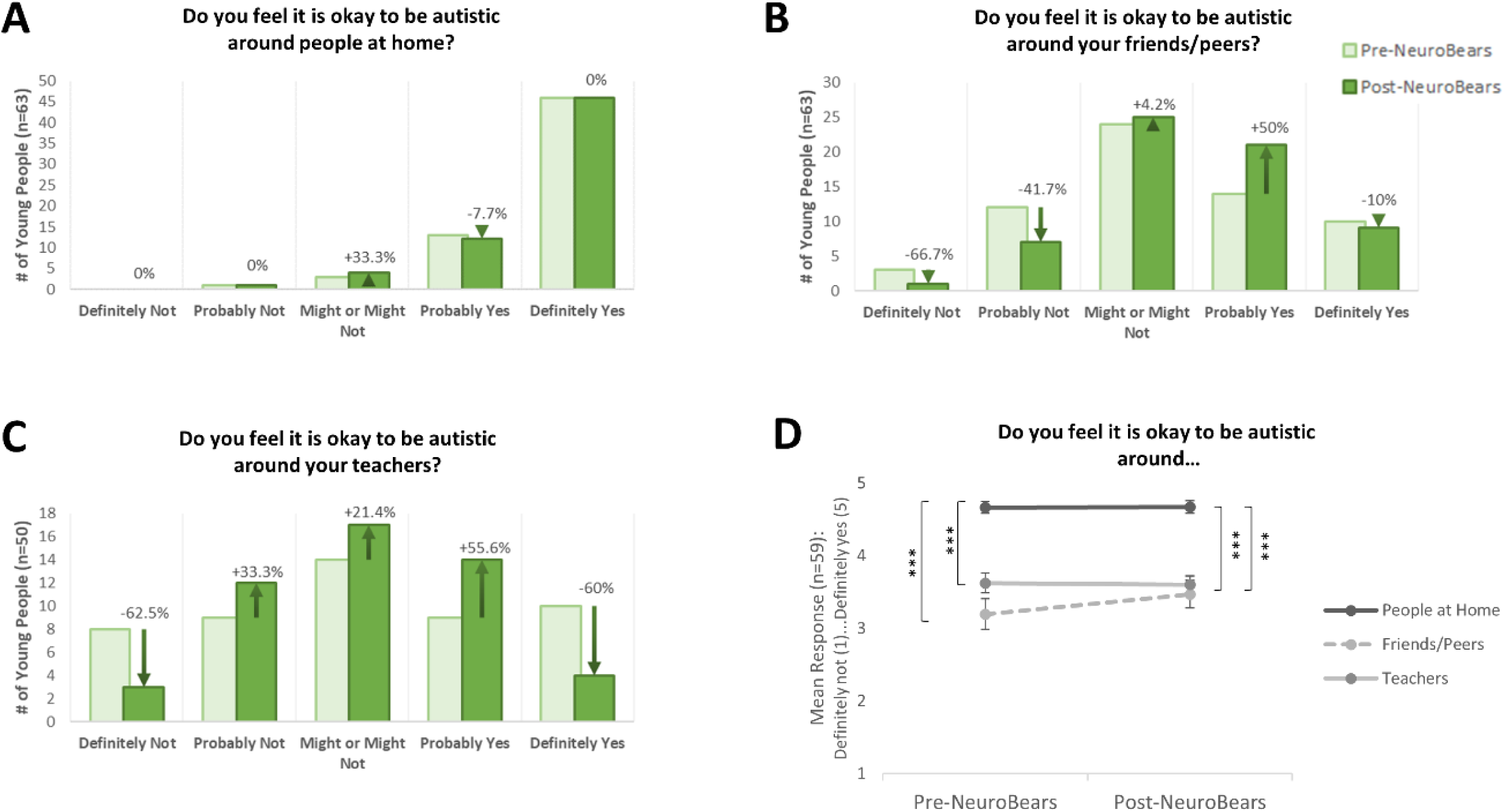
Impact of Context on Feelings about Being Autistic. Children’s responses to the question of whether they feel it is okay to be autistic (A) around people at home, (B) around your friends/peers, and (C) around your teachers, pre- and post-NeuroBears. Whilst no significant change was observed within context pre- and post-NeuroBears (A-C), significant between-contexts were observed whereby the children felt it was significantly less okay to be autistic with both their friends/peers and their teachers, relative to people at home. *** *p* < .001.

Exploring this further, related-samples Friedman’s analysis of variance by ranks revealed a significant difference across the three contexts both pre-NeuroBears (χ^2^ (2) = 41.61, *p* < .001) and post-NeuroBears (χ^2^ (2) = 39.68, *p* < .001), with Dunn-Bonferroni post hoc tests revealing that the children reported feeling significantly more comfortable about being autistic with people at home relative to when they are with their friends/peers (pre-NeuroBears: *p* < .001; post-NeuroBears: *p* < .001) or with their teachers (pre-NeuroBears: *p* < .001; post-NeuroBears: *p* < .001) (Bonferroni adjustments applied). There were no significant changes with respect to how the children reported feeling about being autistic in any of these contexts post-relative to pre-NeuroBears (‘People at Home’: *p* = .952; ‘Friends/Peers’: *p* = .094; ‘with Teachers’: *p* = .834). The absence of change in the ‘People at Home’ context is likely attributable to the fact that there was already a ceiling effect in response to this item Pre-NeuroBears (Figure 12A and 12D). This same pattern was also evident when the children were asked about whether they feel that they can act in a way that feels comfortable and happy for them in three specific contexts ‘At Home’, ‘Out and About’, and ‘At School’ (see Figure S1 for full details).

### 3.5. Non-completion of NeuroBears

Whilst 136 children consented to participate and completed the baseline, pre-NeuroBears questionnaire, only 63 of this group completed both the pre- and post-NeuroBears questionnaire and completed sufficient amount of the NeuroBears course content to be included in the above analysis. An additional 9 children, despite completely very little or none of the NeuroBears course, did complete at least some of the post-NeuroBears questionnaire, and hence provided information on why they didn’t engage with the course. Exploring the reasons provided by these children was informative. For instance, one child reported that their ADHD made it hard for them to stay on track during the course content, and another child stated that “*I didn’t want to do it all because it was too tricky to sit down for a while*”. A third child stated, “*I didn’t want to learn about autism*” and a fourth stated, “*It was tricky because I was not really interested*”. Other reasons included the timing of the research coinciding with holidays or illness, and demand avoidance was referred to by one child.

In addition, we explored whether there were any differences in how these 9 children responded in the pre-NeuroBears questionnaire, relative to the 63 children who did engage with NeuroBears. No significant group differences were found for parameters tapping into the children’s prior understanding of autism. However, the group of children who opted not to engage with NeuroBears, reported feeling significantly more negative about being autistic at baseline (*U* = 162.0, *z* = -2.129, *p* < .05) than their peers who were willing to complete NeuroBears. They were also significantly less likely to talk about autism in their daily lives than their peers who engaged with NeuroBears (*U* = 172.0, *z* = -2.011, *p* < .05), and reported feeling less brave (*U* = 169.0, *z* = -1.988, *p* < .05) and more negative (*U* = 175.5, *z* = -2.213, *p* < .05) when they did talk to others about being autistic.

### 3.6. Created by autistic individuals, for autistic children

When asked “How important is it to you that NeuroBears was created by autistic people?”, 69.5% of children who engaged with NeuroBears stated that it was “Extremely Important” to them and 23.7% reported that it was “Very Important”, whilst 1.7% indicated that it was only “Moderately Important”, 3.4% only “Slightly Important”, and 1.7% “Not at all Important”. In addition, 68.6% of the children also reported that it was either “Extremely helpful” or “Very Helpful” to hear the examples of the creators’ own autistic lived experiences within NeuroBears.

## 4. Discussion

This study demonstrated that an autistic-led and created online psychoeducation course for autistic children was of significant benefit across a range of areas. These included a significant improvement in the children’s knowledge and understanding of being autistic and of their unique strengths and challenges, a significant rebalancing of how the children viewed being autistic, evidence of emerging positive autistic identities and a growing sense of belongingness, a significant change in the children’s abilities to communicate about being autistic, and evidence of strengthening self-advocacy skills.

### Knowledge and Understanding

Following NeuroBears, children reported knowing significantly more about their autistic brains and having significantly greater understanding about how their experience of the world is different to that of a person who is not autistic. The children were also significantly more likely to recognise that they have co-occurring conditions post-NeuroBears.

By asking the children specifically what they had learnt by completing NeuroBears, the children provided valuable insights into what this learning looked like. Some children reflected more on the factual content, e.g., [I learnt]…“*facts about autism*”, “*about autism and different brains*”, “*what is an autistic person and non-autistic person*”, “*about how the autistic brain works*”, “*Lots about being autistic*”, and “*what autism isn’t*”, and that people sometimes have misconceptions about what autism is, e.g., [I learnt]…“*lots of things people think about autistic people that aren’t actually real*”. Children also mentioned learning *“about all the different senses*”, and about concepts such as “*Energy accounting*”, “*masking*”, and regulation, *e.g., “I have learnt a bit more about ways regulation can happen and the concept of it”*.

Some children went a step further, reflecting on how this knowledge led to increased self-understanding, e.g., “[I learnt] *what my autism is and how it affects me*”, “[I learnt] *a lot about how my brain is different to others and I can definitely see the autistic traits I have*”, “[I learnt] *Some of the things I do are because I have autism*”, “[I learnt] *about why i feel things differently to other people and why I find some things hard*”, *“I learnt I have a lot of smells I despise and sounds I despise and how I loose spoons quicker than I thought”, “[I learnt about] my shutdowns”, “I learned the most when thinking about the sensory cars and talked to my mum about how it feels for me”,* and “*I understand why I do things in a certain way*”, whilst other children reflected on specific strategies or benefits that they had taken from NeuroBears, e.g., “[I learnt about] *Having to have lots of notice before going to things*”, “[I learnt about] *things that help with regulation*”, and “*A lot of feeling that i couldn’t visualise were put into analogys which help me recognise what im feeling*”. Moreover, when reflecting on their increased knowledge of co-occurring conditions, one child commented that “*When the course talked about dysgraphia, it really felt like me*”, whilst another stated that the co-occurring conditions section of the course *“made me realise that there was other things too and things we have not spoken about before.”*

Given previous findings that highlight that an important benefit of receiving an autism diagnosis is allowing one to gain an understanding of one’s individual needs (Stagg & Belcher, 2019), and given that many adults diagnosed in adulthood highlight the need for post-diagnostic support to help them gain understanding of what being autistic means/looks like for them personally (26), it is positive to see these autistic children gaining these insights at a young age (8-14 years old, mean age = 10 years). In a previous small and all male sample of autistic youth (aged 9-16), 2 out of the 9 children interviewed described positive aspects of their diagnosis, including an increase in other people’s understanding of them and increased support, however none of the children or their parents described the diagnosis as increasing the child’s self-understanding (2). Indeed, in this study, most of the children were reported to have expressed indifference about their diagnosis, e.g., “*I’m not that interested [in the meaning of the diagnosis]. (YP1)*” (pp. 235), and when the children did attempt to describe the meaning of their diagnoses to the researchers, they defined it in terms of their own primary difficulty, e.g., “*It means you find it hard to make friends because I find it hard to make friends. (YP4)*”. From these and similar statements, the authors concluded that these children lacked a sense of their diagnosis’ broader meaning (2), hence limiting the utility of having the diagnosis in the first instance for these children. Moreover, when asked about what their diagnosis meant, most of the children (78%) in Ruiz Calzada et al., described negative aspects of their diagnosis.

Future studies are needed to explicitly explore the downstream benefits of acquiring this broader, non-deficit focused knowledge of autism and co-occurring conditions in childhood across a wide range of life outcomes. One recent cross-sectional study involving 78 autistic university students provides some initial evidence, finding that being told you are autistic earlier in life was associated with better well-being and quality of life in adulthood (31). The authors here concluded that “*telling a child that they are autistic at a younger age empowers them by providing access to support and a foundation for self-understanding that helps them thrive in adulthood*” (pp. 1).

However, the children in (2) remind us that telling a child of their diagnosis alone is unlikely to be sufficient to achieve these gains, as children likely also need broader knowledge, alongside a positive autistic identity, to counteract negative sense making (3), to benefit in this way. Indeed, the autistic young adults who participated in Oredipe et al. (2023) stressed how young children should learn that they are autistic “in ways that help them understand and feel good about who they are”, and the impact of the negative framing of autism in the diagnostic process should also not be underestimated, e.g., “*Reading the report, it is written in quite a negative way. Because it is the deficit model. So it was quite hard reading the things about myself that I am bad at. Cause I have tried my best to hide them, for the last 40 years”*; “*the diagnosis had a knock-on effect on my confidence at that time [. . .] I think I needed probably a bit of help to process some of that stuff*” (26) (pp. 5).

The children’s descriptions in our study stand in sharp contrast to findings of Crompton et al. (2023) who, like Ruiz Calzada et al. (2012), found that autistic school children often lack an understanding of what being autistic means (32) and that the absence of this knowledge can both deny children the opportunity to share their diagnosis with others, e.g.,

> “*I think the only reason I didn’t tell friends is because I just didn’t know how to. I didn’t really understand it . . . because no one had really explained it to me. You can’t just walk up to someone you have known for three years and go ‘oh by the way I am autistic’, and if they are like ‘oh what does that mean’ say ‘oh I don’t know’. It doesn’t really sound good at all, it is like you are just making it up. So I didn’t know how to say it*.” (Participant 8)

and/or increase stigma when that information is disclosed, e.g.,

> “*they* [my non-autistic peer group] *did know* [that I was autistic], *but I don’t think they understood this. I was still expected to be like them, and communication breakdowns were blamed on me. There was no good information for them, or for me to share. I was just seen as a bit weird*.” (Participant 3).

Interestingly, in our study, one child told us that having this increased knowledge was important for them as it helped them better understand the needs of other neurodivergent people in their lives (“[I learnt] *more about my friends who have ADHD, and what their needs could be*”). Hence, providing autistic children with this wider knowledge might not only support increased self-understanding, but also confer benefits to those around them. This may be particularly important given that autistic children typically grow up in families in which neurodivergences are commonplace.

### Emergent positive identities and autistic solidarity

As described above, positive autistic identities emerge when both the positive aspects of being autistic are recognised alongside the challenges (Bolte, 2023). Indeed, the 20 autistic adults who participated in (21) considered their autism with neutrality and akin to any other personal attribute such as race or hair colour. Completing NeuroBears appeared to rebalance the children’s perception of their autism in terms of the strengths and challenges it brings. More specifically, pre-NeuroBears, less than one-third (30.6%) of the children believed that there are ‘a lot’ or ‘quite a lot’ of things that they are good at because they are autistic, and over half (50.8%) believed that things are ‘quite a lot’ or ‘a lot’ harder for them because they are autistic. This picture reversed post-NeuroBears, with now almost half of the children (46.7%) believing that there are ‘a lot’ or ‘quite a lot’ of things that they are good at because they are autistic, and only a minority of children (14.3%) now reporting believing that things are either ‘a lot’ or ‘quite a lot’ harder for them because they are autistic. This significant rebalancing of their perspectives is likely important in supporting the emergence of positive autistic identities.

Personal strengths that the children recognised in themselves that they linked to their autism included LEGO, gaming, music, maths, art, reading, having a good memory, and solving puzzles, whilst specific challenges included “*talking to people*”, “*Going out, being nervous, having friends*”, “*Because you can be bullied about being different*”, “*Being in the dark and being on my own*”, “*when people don’t keep their promises*”, “*Solving maths equations*”, “*Sensitive (hair can’t be brushed without sensitivity)”, “sometimes I can’t understand why people get annoyed with me about my preferences*”, “*Harder to stop things I regularly do that people don’t want me doing*”, and “*Scholl!!!! School is so hard and changes and when i camt talk and nobody understands how hard it is for me as i cant explain it well enough and i just say im fine at school but im not*”.

Moreover, fundamental to the lived experience of being autistic and to an individuals’ autistic identity, is how you feel about being autistic. Post-NeuroBears, the children’s responses to the question of whether they like being autistic was significantly more positive than pre-NeuroBears, and the children endorsed fewer negative emotions when considering how they feel about being autistic post-NeuroBears relative to pre-NeuroBears. A significant positive relationship was also found between how much the children know about their autistic brains and how much they like being autistic. Autistic adults have previously reported that reframing to a more positive understanding of autism has helped them to manage the constant exposure to stigma that they face, alongside the reclamation of language (i.e., using the word ‘autistic’) and using concealment and disclosure strategically (21). Hence, helping children to understand autism from a neutral, non-pathologized and pragmatic perspective will hopefully support these children to counteract some of the societal stigma surrounding autism that they face, and will continue to face, in life.

This significant and positive change in the children’s feelings about being autistic post-NeuroBears is also notable, given previous findings showing that greater personal autism acceptance has been found to predict lower depressive symptoms in autistic adults (33) and that a positive autistic identity may act as a protective factor against mental health difficulties (18, 20).

In their responses to the question of whether they like being autistic, many of the children spoke movingly about self-acceptance, about no longer feeling so alone, and of a growing awareness of a wider autistic community to which they belong when considering what they learnt through NeuroBears, e.g., “[I learnt] *That it’s ok to be autistic*”, “[I learnt] *That its okay to stim*”, “[I learnt] *That it’s ok to be myself*”, “[I learnt] *That just because your autistic doesn’t mean you’re weird, silly and different-your still human*”, “[I learnt] *it’s hard being autistic but it’s also amazing at the same time*”, “[I learnt] *That’s there’s lots of people like me in the world*”, “[I learnt] *That I M not alone*”, and “[I learnt] *autism isn’t a disorder – I AM autistic, I have a community*”.

These testimonies stand in stark contrast to those documented by Humphreys and Lewis (2008), where the autistic youths they interviewed used language such “a retard” and “a freak” when describing themselves and expressed the desire to be made “normal” in order to fit in (“‘*Sometimes it’s like, “make me normal*”, pp. 31).

Qualitative studies from the adult autism literature suggest that repeated adverse social interactions, particularly early interactions with school peers, cause one to internalise the negative attitudes and reactions of others to them, in turn leading to the formation of negative self-identity and mental health difficulties (34–36). In the developmental literature more widely, peer rejection and the absence of friendship in childhood have both been shown to be associated with psychopathological symptoms in adulthood (37), and loneliness has been found to mediate the relationship between social contact and poor mental health in young autistic adults (Schiltz et al., 2021). Such findings highlight the importance of supporting autistic children to find their neurokin (i.e., their autistic peers), as does the finding of Cooper et al. (2023), which reported that autistic young people had better psychological well-being when they felt more solidarity with other autistic people.

One explanation for why having relationships with autistic peers is beneficial was offered to us by the children in our cohort pre-NeuroBears, when they told us that other neurodivergent children don’t make them feel othered in the way that being the only autistic child in their class at school does, and hence they particularly valued these relationships (Mullally et al., in preparation). Both building connections with like-minded peers, and feeling part of a community post autism diagnosis, have also been described as particularly important by autistic adults, with relationships with other autistic people described as easier and more comfortable than relationships with non-autistic people, and leading to increased feelings of similarity and connection and thus a comfortable and supportive environment, e.g., “*’It just helps to have a group of fellow travellers who understand*’ (26) (pp. 7). Developmental research has previously demonstrated the strong positive impact of friends on children’s overall sense of self-worth (37, 38).

### Masking

Despite the above, and in line with previous findings (3), masking was highly prevalent, and this did not change post-NeuroBears. However, there was a significant relationship between how much the children like being autistic and masking less, suggesting that forming a positive autistic identity may reduce masking in autistic children. This is likely important as autistic individuals who hide their autistic traits show increased risk of experiencing thwarted belongingness and lifetime suicidality (39). Autistic masking has also been linked to autistic burnout (40) and higher rates of depression (33). Hence, heavy reliance on masking in all aspects of life is not optimal for positive mental health.

However, autistic masking also functions as an important safety mechanism (41). When reflecting on the intersection between liking being autistic and trying to hide being autistic observed here, it is important to also consider environment, safe relationships, and agency. As a group, the children identified there were safe spaces and safe people and these were typically home, close family members or neurodivergent peers (42). These were normally framed around feeling understood and feeling safe to unmask (i.e., to be authentically autistic without fear of negative consequences), as has previously been reported in the adult autism literature e.g., “*It is great to have people who don’t need an explanation. . . you don’t need to hide your autistic traits. . .you know there is no need to worry with them*.” (26) (pp. 7).

Within the NeuroBears course, it is recognised that it is not always safe to unmask, and that masking can be used as a tool if you have knowledge and self-understanding (for further discussion see (10, 21, 41)). Reinforcing this, both pre- and post-NeuroBears, the children reported feeling significantly more comfortable about being autistic with people at home, relative to when they are with their friends/peers or with their teachers (Figure 12). They also reported feeling more able to be their true authentic selves at home, but significantly less so when around their friends/peers or when around their teachers (Figure S1), likely due to feeling the need to hide/suppress visible autistic stims (such as hand-flapping) when with peers or teachers due to fear these will identify them as ‘disabled’ or ‘different’, and thus leading others to view them negatively (3).

Given the rates of victimisation of autistic school children, this is unsurprising. More specifically, in a review of 15 studies exploring the prevalence of victimization in autistic school children, autistic children were found to be victimized more frequently than school children in the general population (43, 44), and at alarmingly high rates regardless of whether the rates were reported for weekly (range: 0.4–18%), monthly (range: 7–77%), or yearly (range: 46.3–94%) epochs (45). Moreover, Sreckovic et al. highlighted that autistic students have been found to be victimized more frequently than both their disabled (46, 47) and non-disabled (47, 48) peers. Hence, protecting yourself from bullying via masking autistic traits and ‘difference’, in a society in which autistic traits are seen as a bad traits to have (21), is likely a protective strategy.

More generally, autistic people are at increased risk of societal stigmatization, marginalization, minority stress, discrimination and victimisation (Botha & Cage, 2022; Butler & Gillis, 2011; Obeid et al., 2015; Pearson et al., 2023; Wood & Freeth, 2016), and autistic vulnerability to negative life events such as victimisation has been found to partially mediate the relationship between autism and mental health difficulties (Griffiths et al., 2019), which are highly prevalent in autistic adults (49, 50) and children (Dickerson Mayes et al., 2015) alike. Gaining the confidence to self-advocate for one’s needs may offer important protection against events such victimisation. Moreover, developing a robust positive identity and related positive self-esteem (20) likely confers protective benefits against the impact of bullying, stigma, and victimisation (51). Hence, helping autistic children to develop skills that could protect them against the widespread victimisation that they face (45), and/or insulate them somewhat from the psychological impact of traumatic events (52), should be prioritised. Exploring the role that psychoeducation autism courses (such as NeuroBears) could play in facilitating the development of such skills, is clearly important.

### Communication and self-advocacy

Essential to self-advocacy is being able to communicate your specific needs. Post-NeuroBears, the children reported feeling significantly braver to tell other people what they need, and this increased sense of safety was explicitly linked by one child to having learnt more about themselves and autism through NeuroBears: “*I felt safe as I learnt more about myself and autism*”.

A fundamental steppingstone to self-advocacy (and indeed to self-understanding) is having the language to communicate about your autistic experiences. Post-NeuroBears, significantly more children reported having this language. They also reported communicating with family about their experiences significantly more frequently, and that they felt significantly more positive about approaching these discussions. Extending this further, one child stated: “*I feel a bit more confident talking about being autistic since watching NeuroBears*”.

Despite these positive changes, conversations about being autistic remained challenging and the children frequently reporting feeling anxious in these situations, with communication with others outside of the family being particularly challenging even post-NeuroBears. This was particularly true for the children who had the least prior knowledge of autism at baseline (see below for further discussion). Despite these challenges, there were concrete examples of how the newly acquired knowledge and language was being used by the children to self-advocate post-NeuroBears, e.g., “*I made a power point about autism and showed it to my teacher so he could understand me better*”. Future research is needed to establish whether such benefits to communication and emergent self-advocacy transfer into better wellbeing downstream.

NeuroBears recognises that children often have little control over their environments and relationships. However, knowledge of autism, regulation, masking, and wider autistic culture can lead to recognition of safe spaces, environments, and people, hence, NeuroBears encourages children to seek these out where possible. NeuroBears hopes that giving autistic children this knowledge can help them to more safely navigate the often complex and dangerous terrains autistic people face in everyday life (41, 45) and thus, in so far as possible, to shape their own narratives.

### Prior Knowledge

Children commenced the research with varying levels of pre-existing knowledge about autism, ranging from ‘I know nothing at all’ to ‘I know a great deal’. An important aspect of the analysis was to explore the impact of baseline autism knowledge on the above outcomes. For several parameters relating to understanding autism, and communicating about being autistic, the children who started the course with least prior knowledge of autism lagged behind those who entered the research with more prior knowledge.

For some specific items (i.e., the items that measured autistic knowledge, language to talk about being autistic, and how comfortable the children feel when talking to their families about their autistic experiences), the gap between those who started the course with less prior knowledge of autism and those who had more knowledge at baseline, narrowed significantly between the pre- and post-NeuroBears phases (see Figure 4B, 6B, 6F).

For other items, however, this narrowing was either not evident or failed to reach statistical significance. These latter items included items that assessed understanding of autistic experience (Figure 4D), how frequently the children talk about being autistic (Figure 6D), and how comfortable (Figure 7D), brave (Figure 9B) and safe (Figure 9D) the children reported feeling when talking to others outside of the family about their autistic experiences (see also Figure 8B).

These latter items tap into particularly challenging things for all autistic children to do, so it is likely that readiness and confidence in one’s abilities to do these things may take time to develop; time which those who begun the research with greater knowledge of autism have had more of. This resonates with the findings from the adult literature showing that the length of time that has elapsed since diagnosis is associated with having a more positive autistic identity (19), as integrating new knowledge and understanding into one’s personal identity is likely a dynamic, iterative and on-going process (53). Future longitudinal studies are required to assess this formally. Overall, however, the data suggest that children with the least pre-existing knowledge of autism derive considerable benefit from NeuroBears, even if they did not always entirely close the gap with their peers who entered the research knowing more about autism.

### Importance of being created by autistic people

A unique aspect of NeuroBears as a psychoeducation course for autistic children is that it was created, written, and delivered by autistic people. This was rated as being either extremely (69.5%) or very (23.7%) important to the young people. Reasons provided by the children as to why this was the case included: “*It was because they understand because they are the same as me*”, “*Autistic people know what they are talking about when talking about autism*”, and “*Seeing someone like me do something helpful*”. The children also valued hearing examples of the creators’ own autistic lived experiences as part of NeuroBears, with one child commenting “*It was interesting to learn about the stuff about how others deal and experience autism”,* and another “*it didnt make me feel alone*”.

Increasing more positive representations of autistic people in the media has previously been identified as an important way to reduce autism stigma, as autism stigma is typically driven by peoples’ misunderstandings of autism (51). Hence, having autistic individuals create and produce NeuroBears offers the children positive autistic role models right from the offset. This may reduce internalised stigma that they already carry with respect to their autistic identity (21, 27, 54), hence protecting self-concept and wellbeing (54). There is a growing body of research evidencing the potential benefits of autistic-autistic interactions on autistic people’s well-being and sense of belonging, e.g., (26, 32). Whilst this research adds to this, it also flags that autistic (adult)-autistic (child) support is highly valued by autistic children.

### Filling a gap in post-diagnosis support

Despite repeated calls for the provision of sustainable and funded post-diagnostic support for autistic individuals (24–26, 55), such support is rarely available within health services in the UK, and what is available is generally considered inadequate.

Underlining this are the findings of Rodgers et al. (2016), who surveyed 116 UK multidisciplinary professionals clinically active in autism diagnosis and assessment services. Professionals included psychologists, speech and language therapists, paediatricians, and psychiatrists predominantly working within the National Health Service (NHS). This study found that the most frequently offered post-diagnostic supports were “Information on support groups” (‘always’ offered in 65% of services) and “Information leaflets” (‘always’ offered in 64% of services). “Education/support group for parents” was only ‘always’ offered in 26% of services, and an “education/support group for patients” was only ‘always’ offered in 9.5% of services. Moreover, less than half of the professionals reported feeling satisfied with in-service post-diagnostic provision - with particular concern about post-diagnostic support for primary school age children and young adults.

Parents of autistic children also frequently raise the inadequacy of current post-diagnostic support, e.g., “[we needed] *more information of where you can get help rather than just sort of…be dumped after the diagnosis*” (24) (pp. 3767), “*After the very considerate diagnostic process and level of care, we were left in the dark. We were given no information … a few leaflets*” (56) (pp. 159) and “*it was out the door and I was on my own”* (25) (pp. 761).

Critical, however, is the lack of support in understanding their diagnosis, fear, and/or a loss of self-esteem that autistic individuals have described feeling post-diagnosis e.g., “There wasn’t any help” and “*it was quite. . . well it was frustrating but it also it was quite. . . in a way quite scary to be sort of. . . left unsure how to manage this in. . . from the rest of my life going forward*” (26) (pp. 5).

A ten-week autistic-led programme (‘Exploring Being Autistic’ https://www.autismmatters.org.uk/exploring-being-autistic.html), aimed at autism psychoeducation within a peer group context, has been successfully trialled in two groups of 16 recently diagnosed/self-identified adults, both in an in-person (57) (mean age of participants = 44.2 years) and online (58) (mean age of participants = 49.2 years)) format. Thematic analysis revealed that the autistic-led nature of the programme was appreciated by the autistic adults, as was developing a positive, practical outlook on autism. However, this course was developed to support autistic adults (aged 19-74 years), with participants typically aged between 30 and 50. For all the reasons described above, gaining such understanding and sense of community as early in life as possible, is likely important.

Moreover, whilst social media groups have successfully supported members of the autistic community to gain understanding of their diagnosis (Crompton et al., 2022), social media should never be what plugs a gap in health care provision. This is even more pertinent for young children and vulnerable youth. Indeed, even the adult participants in Crompton et al. (2022) reported struggling with social media groups and described the negative impact these can have, e.g., “*Some people are very angry on there [social media] or really struggling and sometimes it can be quite depressing to read those things*”. Heath service commissioners and service providers need to do better for their service users.

Importantly, NeuroBears also supports the children’s safe adult(s) throughout, developing their knowledge of autism, common autistic experiences and co-occurring conditions. In doing so, it seeks to build the safe adult’s confidence talking to their child about their autistic experiences, and hence further enable the emergence of open dialogue between the children and their safe adult(s) and provide both the child and adult with the skills to more knowledgeably advocate for themselves.

Whilst data from the safe adults was also collected as part of this research, it will be published elsewhere. In this research, all safe adults were the children’s parent(s) and hence NeuroBears supported both the child and the parent(s).

### Strengths and Limitations

Strengths of this research include the large sample size and relatively good representation of sex, which is often skewed in favour of male participants in autism research. However, the sample did not include any children with an intellectual disability and was not representative in terms of socio-economic status or ethnicity. The reasons for this are likely complex and discussed in further detail elsewhere (Mullally et al., in preparation). The sample was also restricted to individuals currently residing in the United Kingdom.

Another issue related to recruitment that was initially encountered was that, due to study recruitment occurring online, we had a larger than anticipated group of autistic children who entered the research with an already strong knowledge of autism, potentially leading to ceiling effects in the research. We addressed this potential issue at recruitment by actively encouraging sign-up from families new to autism. This resulted in us having enough participants with a broad range of baseline knowledge to enable the subdivision into three groups and inclusion of this as a variable in our analyses.

We also encountered a pattern with the question format for some questions, whereby general, less specified question formats were less successful at detecting change than more specified questions. Such questions also had less specified response options, so it is unclear whether it was the more general question format or more general response options that may have been an issue. For instance, when asked “Do you talk about being autistic” [Response options: Yes a lot, Quite a lot, Somewhat, A little, Never], we failed to detect a significant change pre- and post-NeuroBears (*p* = .062), however when this question was re-phrased in a more specific manner, made specifically relevant to conversations with family [“How often do you talk to others (e.g., family) about your autistic experience?”], and had more concrete response options [Everyday, 5-6 days a week, 3-4 days a week, 1-2 days a week, Never], a significant change was evident (*p* = .009). The children themselves also told us that they sometime struggled with the less specified questions and hence, moving forward, these findings should be considered when designing questionnaires and semi-structured interviews for autistic children.

We also had several families withdraw from the research due to the children’s demand avoidant profiles making it difficult for them to engage with the research, and despite working with families to provide accommodations, for some children, it simply was not the right time for them to engage with the research. This was also the case for some children who were experiencing mental health difficulties. The 6-week time limit to complete NeuroBears was also not ideal for some families, particularly if there were other co-occurring life events (such as illness or holidays e.g., “*I was very overwhelmed by Christmas and getting through all of that, so I didn’t want to have anything extra to do. I’m going to try it over the summer instead*”), and this increased the attrition rate between phases. This time limit was necessary due to the research design, but this would not be applicable outside of the research context. As described above, other children reported having difficulties staying on track or sitting down to engage. Additional consideration should be given to how to support engagement in these children, perhaps via simplified bite-sized video content.

As all participants, regardless of whether they completed NeuroBears or not, were invited to complete both the pre- and post-NeuroBears questionnaires, we were able to explore whether there were any notable differences at baseline (i.e., pre-NeuroBears) between the children who reported engaging with NeuroBears (n=63) and those who told us they did not (n=9). Those who did not engage, initially reported feeling significantly more negative about being autistic, reported talking to others about being autistic significantly less frequently, and felt significantly more negative and significantly less brave when they did have these conversations. Hence, it is probable that these internalised negative views of autism and the difficult emotions experienced when discussing being autistic played a role in these children’s reluctance to engage with an autism psychoeducation course. A similar possibility was proposed by Ruiz Calzada et al. (2012) when they noted that the autistic young people that they spoke with expressed disinterest in learning about their diagnoses but also held inherently negative views of autism. Overcoming this reluctance to engage with learning about autism is important but attempts to do so need to recognise where this reluctance may be stemming from – and hence approached gradually, carefully and in a supportive manner.

Challenging stigmatizing and deficit-based autism narratives that the child may have attuned to, may help create a safe space for the child to be able to engage in discussions about being autistic. Future research is needed to better understand how this could be achieved. In the meantime, it is important to consider the voice of one of our participants who told us “*I am glad NeuroBears is there for when I decide I’m interested to know more*”.

Finally, the post-NeuroBears questionnaire was completed shortly after completing the NeuroBears course. Further longitudinal research is required to assess for downstream impact.

## Conclusion

Diagnostic criteria seek to condense individual information into a circumscribed entity, and in doing so “risk that an individual is perceived primarily within these confines” (Bölte et al., 2022, pp. 3), potentially not just by others, but also by themselves. After 120 years of autism clinical practice (59), we are still not adequately supporting autistic people. NeuroBears takes a different approach, closely aligned to the social model of disability and informed by the autistic lived experience. In NeuroBears, children learn about being autistic in a neutral and non-stigmatizing manner, and in a way that does not confine them to a list of symptoms or deficits. Rather than a top-down system, where autistic children can be mis-interpreted or mis-represented, the autistic community favours building a bottom-up system that empowers the child to build their own autistic identity via authentic self-understanding. NeuroBears embodies this approach, and here we showed the numerous benefits that this has for autistic children’s self-understanding, emergent identity, sense of belonging and self-advocacy skills.

## Supporting information

see Supplemental Materials (pages 2-4).

## Data Availability

All data produced in the present study are available upon reasonable request to the authors

## Acknowledgments

We sincerely thank all the children and families who generously gave up their time to take part in this research and to share their stories with us. We wish them all the very best in the future. We also acknowledge Nic King (www.pandasonline.org) for her support and contributions to the research.

