## Supplementary material for "“I like being autistic”: Assessing the benefit of autistic-led psychoeducation for autistic children": see Supplemental Materials (pages 2-4).

### Pre- and Post-NeuroBears - Questions and Response Options

1. ***Understanding of autism*:**
   i. **How much do you know about your autistic brain?** [Response Options: 'I know a great deal' (5), 'I know a lot' (4), 'I know somethings' (3), 'I know a little' (2), 'I know nothing' (1)].

   ii. **How much do you understand about how your experience of the world is different to a person who is not autistic?** [Response Options: 'I understand a great deal' (5), 'I understand lot' (4), 'I understand somethings' (3), 'I understand a little' (2), 'I understand nothing' (1)].

   iii. **Do you think that you have any co-occurring/other conditions, as well as being autistic?** [Response Options: 'Definitely Yes' (5), 'Probably Yes' (4), 'Maybe' (3), 'Probably No' (2), 'Definitely No' (1)].

   iv. **Do you think there are any good things about being autistic?** [Response Options: 'Yes, there are a lot' (5), 'Yes, there are quite a lot' (4), 'Yes, there are some' (3), 'Yes, but only a few things' (2), 'No nothing' (1)].

   v. **Do you feel you are good at some things because you are autistic?** [Response Options: 'Yes, there are a lot' (5), 'Yes, there are quite a lot' (4), 'Yes, there are some' (3), 'Yes, but only a few things' (2), 'No nothing' (1)].

   vi. **Do you feel some things are harder for you because you are autistic?** [Response Options: 'Yes, there are a lot' (5), 'Yes, there are quite a lot' (4), 'Yes, there are some' (3), 'Yes, but only a few things' (2), 'No nothing' (1)].

2. ***Communicating about being autistic (Pre- and Post-NeuroBears)*:**
   i. **Do you talk about being autistic?** [Response Options: 'Yes a lot' (5), 'Quite a lot' (4), 'Somewhat' (3), 'A little' (2), 'Never' (1)].

   ii. **How often do you talk to others (e.g., family) about your autistic experience?** [Response Options: 'Everyday' (5), '5-6 days a week' (4), '3-4 days a week' (3), '1-2 days a week' (2), 'Never' (1)].

   iii. **Do you feel you have the words to talk about being autistic? [**Response Options: 'Definitely Yes' (5), 'Probably Yes' (4), 'Maybe' (3), 'Probably No' (2), 'Definitely No' (1)].

   iv. **How comfortable do you feel talking about being autistic with your family? [**Response Options: 'Extremely comfortable' (5), 'Somewhat comfortable' (4), 'Neither comfortable nor uncomfortable' (3), 'Somewhat uncomfortable' (2), 'Extremely uncomfortable' (1)].

   v. **How comfortable do you feel talking about being autistic with other people (other than your family)? [**Response Options: 'Extremely comfortable' (5), 'Somewhat comfortable' (4), 'Neither comfortable nor uncomfortable' (3), 'Somewhat uncomfortable' (2), 'Extremely uncomfortable' (1)].

vi. **How do you feel when talking to other people about being autistic?** [Response Options (select all that apply): Happy, Comfortable, Calm, Excited, Accepting, Engaged, Okay, Numb, Tired, Sad, Angry, Frustrated, Scared, Anxious, Depressed, Unwell, Panicked, Nervous].

*vii.* **How brave do you feel to talk about being autistic?** [Response Options: 'Very' (5), 'Mostly' (4), 'Somewhat' (3), 'A little' (2), 'Not at all' (1)].

viii. **How often do you feel safe to or able to talk to others (e.g., family) about your autistic experience?** [Response Options: 'Everyday' (5), '5-6 days a week' (4), '3-4 days a week' (3), '1-2 days a week' (2), 'Never' (1)].

##### 3. ***Self-Advocacy (Pre- and Post-NeuroBears)*:**

i. **How brave do you feel to tell other people what you need?** [Response Options: 'I know a great deal' (5), 'I know a lot' (4), 'I know somethings' (3), 'I know a little' (2), 'I know nothing' (1).

ii. **Do you always feel brave enough to tell others what you need?** [Response Options: 'Extremely comfortable' (5), 'Somewhat comfortable' (4), 'Neither comfortable nor uncomfortable' (3), 'Somewhat uncomfortable' (2), 'Extremely uncomfortable' (1)].

##### 4. ***Feelings about being autistic (Pre- and Post-NeuroBears)*:**

i. **I like being autistic.** [Response Options: 'Strongly agree' (5), 'Agree' (4), 'Neither agree nor disagree' (3), 'Disagree' (2), 'Strongly disagree' (1)].

ii. **I try to hide my autism.** [Response Options: 'Strongly disagree' (5), 'Disagree' (4), 'Neither agree nor disagree' (3), 'Agree' (2), 'Strongly agree' (1)].

iii. **How do you feel about being autistic?** [Response Options (select all that apply): Happy, Comfortable, Calm, Excited, Accepting, Engaged, Okay, Numb, Tired, Sad, Angry, Frustrated, Scared, Anxious, Depressed, Unwell, Panicked, Nervous].

iv. **Do you feel it is okay to be autistic around people at home? [**Response Options: 'Definitely Yes' (5), 'Probably Yes' (4), 'Maybe' (3), 'Probably No' (2), 'Definitely No' (1)].

v. **Do you feel it is okay to be autistic around your friends/peers? [**Response Options: 'Definitely Yes' (5), 'Probably Yes' (4), 'Maybe' (3), 'Probably No' (2), 'Definitely No' (1)].

vi. **Do you feel it is okay to be autistic around your teacher? [**Response Options: 'Definitely Yes' (5), 'Probably Yes' (4), 'Maybe' (3), 'Probably No' (2), 'Definitely No' (1)].

vii. **Do you feel you can act in a way that feels comfortable and happy for you when you're at home?** [Response Options: 'All of the time' (5), 'Most of the time' (4), 'About half of the time' (3), 'Some of the time' (2), 'Never' (1)].

viii. **Do you feel you can act in a way that feels comfortable and happy for you when you're out and about?** [Response Options: 'All of the time' (5), 'Most of the time' (4), 'About half of the time' (3), 'Some of the time' (2), 'Never' (1)].

ix. **Do you feel you can act in a way that feels comfortable and happy for you when you're at school?** [Response Options: 'All of the time' (5), 'Most of the time' (4), 'About half of the time' (3), 'Some of the time' (2), 'Never' (1)].

#### Post-NeuroBears Only - Questions and Response Options:

**1. *Created by autistic individuals, for autistic children (Post-NeuroBears Only)*:**

i. **"How important is it to you that NeuroBears was created by autistic people?"** ['Extremely important' (5), 'Very important' (4), 'Moderately important' (3), 'Slightly important' (2), 'Not at all important' (1)]. [Free text response box].

ii. **"How helpful was it to hear [the creators] examples as part of [NeuroBears]?"** ['Extremely helpful' (5), 'Very helpful' (4), 'Moderately helpful' (3), 'Slightly helpful' (2), 'Not at all helpful' (1)]. [Free text response box].

**2. *Learning (Post-NeuroBears Only)*:**

**i.** **What did you learn during NeuroBears?** [Free text response box].

**Figure S1**

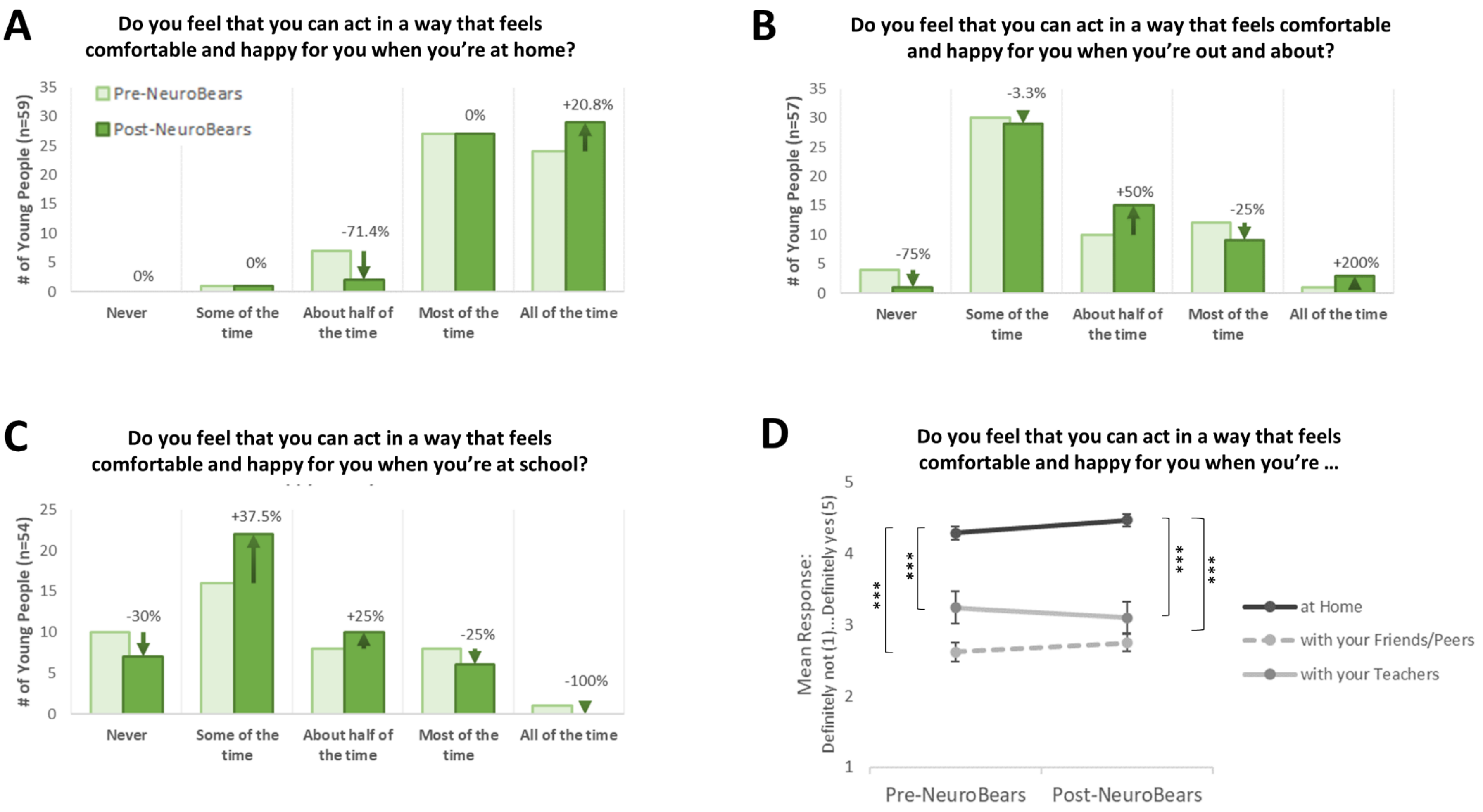
A
Do you feel that you can act in a way that feels comfortable and happy for you when you're at home?
### of Young People (n=59)
Pre-NeuroBears
Post-NeuroBears
0%
0%
-71.4%
0%
+20.8%
Never
Some of the time
About half of the time
Most of the time
All of the time
B
Do you feel that you can act in a way that feels comfortable and happy for you when you're out and about?
### of Young People (n=57)
-75%
-3.3%
+50%
-25%
+200%
C
Do you feel that you can act in a way that feels comfortable and happy for you when you're at school?
### of Young People (n=54)
-30%
+37.5%
+25%
-25%
-100%
D
Do you feel that you can act in a way that feels comfortable and happy for you when you're ...
Mean Response:
Definitely not (1)...Definitely yes (5)
***
at Home
with your Friends/Peers
with your Teachers
Pre-NeuroBears
Post-NeuroBears

**Table S1**

| Correlations: Pre-NeuroBears | | | How much do you know about your autistic brain? | I like being autistic. | I try to hide my autism. |
|---|---|---|---|---|---|
| Spearman's rho | How much do you know about your autistic brain? | Correlation Coefficient | -- | | |
| | | Sig. (2-tailed) | . | | |
| | | N | 63 | | |
| | I like being autistic. | Correlation Coefficient | .257* | -- | |
| | | Sig. (2-tailed) | .042 | . | |
| | | N | 63 | 63 | |
| | I try to hide my autism. | Correlation Coefficient | .038 | -.364** | -- |
| | | Sig. (2-tailed) | .768 | .003 | . |
| | | N | 63 | 63 | 63 |

*. Correlation is significant at the 0.05 level (2-tailed).

**. Correlation is significant at the 0.01 level (2-tailed).
